# The Molecular Endotypes of Type 1 and Type 2 SLE

**DOI:** 10.1101/2022.11.19.22282527

**Authors:** Robert D. Robl, Amanda M. Eudy, Prathyusha S. Bachali, Jennifer L. Rogers, Megan E.B. Clowse, David S. Pisetsky, Peter E. Lipsky

## Abstract

**Objective:** To characterize the molecular landscape of patients with Type 1 and Type 2 systemic SLE erythematosus (SLE) by analyzing gene expression profiles from peripheral blood.

**Methods:** Full transcriptomic RNA sequencing was carried out on whole blood samples from 18 subjects with SLE selected by the presence of manifestations typical of Type 1 and Type 2 SLE. The top 5,000 row variance genes were analyzed by Multiscale Embedded Gene Co-expression Network Analysis (MEGENA) to generate gene co-expression modules, that were functionally annotated and correlated to various demographic traits, clinical features and laboratory measures.

**Results:** Expression of specific gene coexpression modules correlated with individual features of Type 1 and 2 SLE and also effectively segregated samples from Type 1 from Type 2 SLE patients. Unique Type 1 SLE enrichments included IFN, monocytes, T cells, cell cycle, and neurotransmitter pathways, whereas unique Type 2 SLE enrichments included B cells and metabolic and neuromuscular pathways. Gene co-expression modules of Type 2 SLE patients were identified in subsets of previously reported patients with inactive SLE and idiopathic fibromyalgia (FM) and also identified subsets of patients with active SLE with a greater frequency of severe fatigue.

**Conclusion:** Gene co-expression analysis successfully identified unique transcriptional patterns that segregate Type 1 SLE from Type 2 SLE and further identified Type 2 molecular features in patients with inactive SLE or FM and with active SLE with severe fatigue.

## INTRODUCTION

Systemic SLE erythematosus (SLE) is a prototypic autoimmune disease characterized by diverse clinical manifestations that vary in severity and intensity over time (1). Although deposition of immune complexes and the actions of Type 1 interferon can account for at least some manifestations of SLE, many of the symptoms that trouble patients the most, including fatigue and widespread pain have an uncertain relationship to inflammation and immunologic disturbance. Despite their frequency and impact on patients with SLE, these symptoms are not included in criteria for disease classification and are not represented in most measures of disease activity (2).

A new conceptual framework for assessing SLE, that includes pain and fatigue, has been proposed (3). In this model, Type 1 features, such as nephritis, arthritis and cutaneous SLE, are typically inflammatory in origin and can be associated with specific autoantibodies (e.g., anti-DNA and nephritis). In contrast, Type 2 manifestations include widespread pain, fatigue, depression, sleep disturbance and other neuropsychological findings such as “brain fog.” Because of the high frequency of these symptoms in SLE compared to the normal population (4) it has further been posited that Type 2 features are intrinsic features of SLE and related to underlying pathogenesis, even if they might not track with inflammation. It is important to emphasize that signs and symptoms of SLE vary with time and treatment in individual patients and those presenting with Type 1 SLE may evolve into Type 2 and vice versa and those with Type 2 may have persistent or intermittent symptoms (3).

Here, we have used a molecular approach to distinguish Type 1 and Type 2 SLE, testing the hypothesis that the two phases of SLE might arise from distinct pathogenetic disturbances that can be revealed by analysis of gene expression profiles in peripheral blood cells. For this purpose, we used a “bookend” approach and identified patients with isolated Type 1 and Type 2 SLE. The data indicate that patients with Type 1 and Type 2 SLE can be distinguished by analysis of peripheral blood cell gene expression, with the pathways identified providing insights into the mechanisms of these manifestations and potentially pointing to new treatment targets.

## Materials & Methods

### Patient Population

All patients were enrolled in the Duke SLE Registry (DLR) and were adults (≥18 years old) who met 1997 ACR or 2012 SLICC criteria for SLE (5,6). All patients signed informed consent to participate in the registry and for collection of the RNA samples (Duke Health IRB Pro00008875). This was a cross-sectional analysis on a selected subset of 18 patients (Duke Health IRB Pro00094645) using a “bookend” approach that specifically identified patients who had predominant Type 1 or Type 2 disease at the time of analysis. To be included in the Type 1 SLE group, patients had a clinical SLEDAI ≥4, active nephritis, SLEDAI ≥6, or Type 1 Physician Global Assessment (PGA) ≥1 and inactive Type 2 SLE (defined as a Polysymptomatic Distress Scale (PSD) ≤6 and Type 2 PGA ≤0.25). Type 2 SLE patients had active Type 2 SLE symptoms (defined as PSD ≥11 and Type 2 PGA ≥1) and inactive Type 1 SLE (defined as SLEDAI = 0 and Type 1 PGA ≤0.5). Patient identifiers have been completely anonymized and the original patient IDs are completely unknown outside of our research group.

### Data Collection

At the time blood was obtained for gene expression analysis, patients completed the PSD, which includes two subscales: the widespread pain index (WPI) and symptom severity score (SSS) (7–10). In addition to patient-reported measures, patients’ treating rheumatologists completed disease activity measures, including the SLEDAI, PGA for Type 1 activity, and a PGA for Type 2 activity (2,11,12); rheumatologists scored the severity of Type 1 and Type 2 SLE activity separately on scales from 0 (no activity) to 3 (severe activity). (**Supplementary Tables 1 and 2**).

### Gene expression data and gene filtering

Whole blood was collected in PAXgene Blood RNA tubes. After removal of ribosomal RNA and globin transcripts with the Ribo-Zero Globin Removal kit (Illumina), stranded libraries were prepared with the TruSeq Library prep kit (Illumina) and hybridized to a flow cell for sequencing with the Illumina HiSeq platform. The top 5,000 row variances (top5k rowVar) genes determined using standard deviation between samples were retained for further analysis. Data were analyzed for differentially expressed genes (DEGs), for subset clustering by Principal Component Analysis (PCA), and for co-expressed genes using Multiscale Embedded Gene Co-expression Network Analysis (MEGENA) (13) as described in **Supplementary Methods**. Gene expression data from FM patients was obtained from GSE67311 (14) and analyzed as described in the **Supplementary Methods**. Gene expression data from inactive SLE (SLEDAI<6) patients was obtained from GSE45291 (15) and GSE49454 (16). Gene expression data from active SLE patients was obtained from GSE88884 (Illuminate 2). Raw data files have been deposited in NCBI accession PRJNA858861.

## RESULTS

### Patients

Patients had been diagnosed with SLE for a mean of 15.8 years (SD: 7.3) and 55% had a history of SLE nephritis. Seventeen patients were female and one was male; the mean patient age was 41 (**Supplementary fig S1.A)**.

### PCA Groups Type 1 and Type 2 SLE Patients

Initially, we determined that differential gene expression analysis generated only one significant DEG, likely because of the high variance patterns within the two sample sets rather than between them. Therefore, additional analytic approaches were applied to the top5k rowVar genes encoding known proteins. PCA generally separated samples from Type 1 and Type 2 SLE, although 3 outliers were clearly noted (patient IDs Type1_275, Type2_008, and Type2_267 (red arrows, **Supplementary Figure S1 B**). To obtain a preliminary idea of the clinical features segregating with the samples in PCA space, the first four principal component (PC) vectors were correlated to the various recorded sample traits and the top 20 positive or negative correlations per PCA visualized (**Supplementary Figure S1 C**). Of note, PC1 highly correlated to anti-dsDNA, belimumab, and MMF usage, PC2 to NSAID usage, African ancestry, anti-Smith, and anti-RNP, PC3 to cyclophosphamide and amitriptyline usage, and PC4 to PSD score and total areas of pain. These results suggest that Type 1 and Type 2 SLE are largely but not completely separable based on gene expression variance, and that specific clinical characteristics segregate with gene expression variance patterns.

### Gene Co-expression Analysis Identifies Distinct Type 1 and Type 2 Gene Modules

Gene Co-expression analysis was next employed to delineate transcriptomic differences between Type 1 and Type 2 SLE in greater detail. MEGENA, an analytic technique not previously employed with samples from SLE patients, was used to generate co-expression modules from the top5k rowVar genes of the SLE samples (**Supplementary Figure S2**).

To determine the correlation of co-expression modules with clinical features, the module eigengene (ME) of each module was calculated and correlated to the various recorded clinical and demographic traits (**Supplementary Table 3**). Associations between MEs of specific co-expression modules and select clinical and demographic features are shown in **Figure 1 A-I**. The functional nature of co-expression modules was identified by examining genes in each module for overlap with gene modules identifying specific cells or functions in **Figure 1 J-L**.

**Figure 1.**
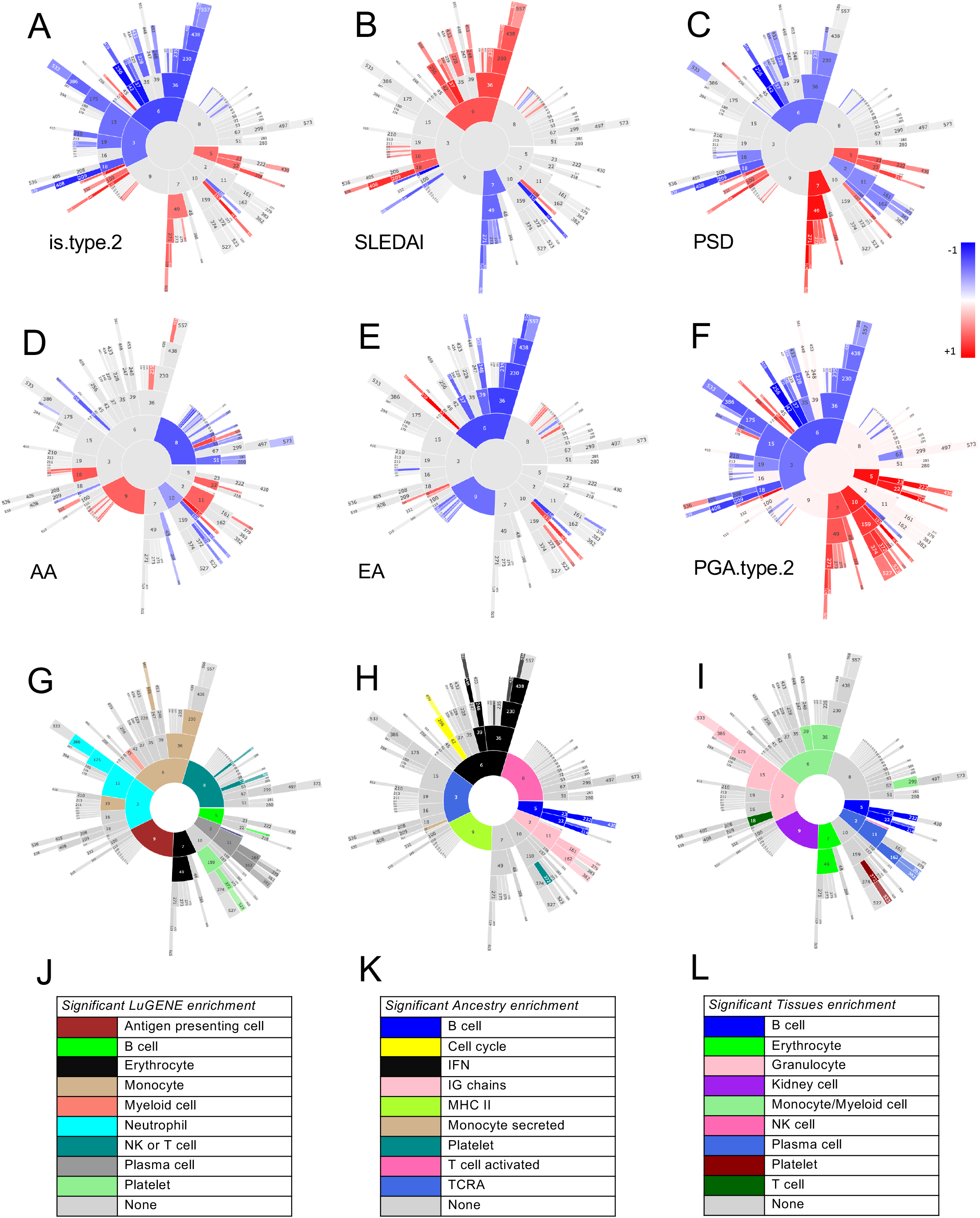
Correlations of MEGENA module expression and various clinical and demographic features. The module eigengene (ME, equivalent to the first principal component) for each module was calculated and Pearson correlations to MEs calculated for multiple demographic and clinical features with correlations ranging from −1 to +1 (A-I). Functional identity of the modules was carried out by matching module genes with various cell type or biological pathway markers (J-L). Functional designation required a minimum overlap of >=3 gene symbols with an associated Fisher’s exact test (p <0.2) to discard overlaps that occurred because of random chance.

The top 40 positive or negative ME correlations correlated to Type 1/2 SLE cohort were identified and submitted to table k-means clustering that revealed groupings of clinical traits and correlated molecular functions (**Figure 2**). Most Type 2 features, including PSD score, PGA Type 2, wake unrefreshed, WPI score, and tired among others were found in the first vertical patient column cluster 1, whereas Type 1 features, including SLEDAI, anti-dsDNA, proteinuria, and EULAR score among others were found in patient cluster 2. Patient cluster 1 showed strong positive correlations to the horizontal module clusters A, E, and G, containing various metabolic pathway signatures and B cells. The Type 1 patient cluster 2 showed strong positive correlations to horizontal module clusters B, C, D, and F signatures including monocytes, interferon, T cells, neutrophils, cell cycle, and other signatures. An alternative depiction of the top 40 intracorrelations is provided in **Supplementary Figure S3**, and correlation pairs of select patient clinical scores and molecular assays in **Supplementary Figure S4**.

**Figure 2.**
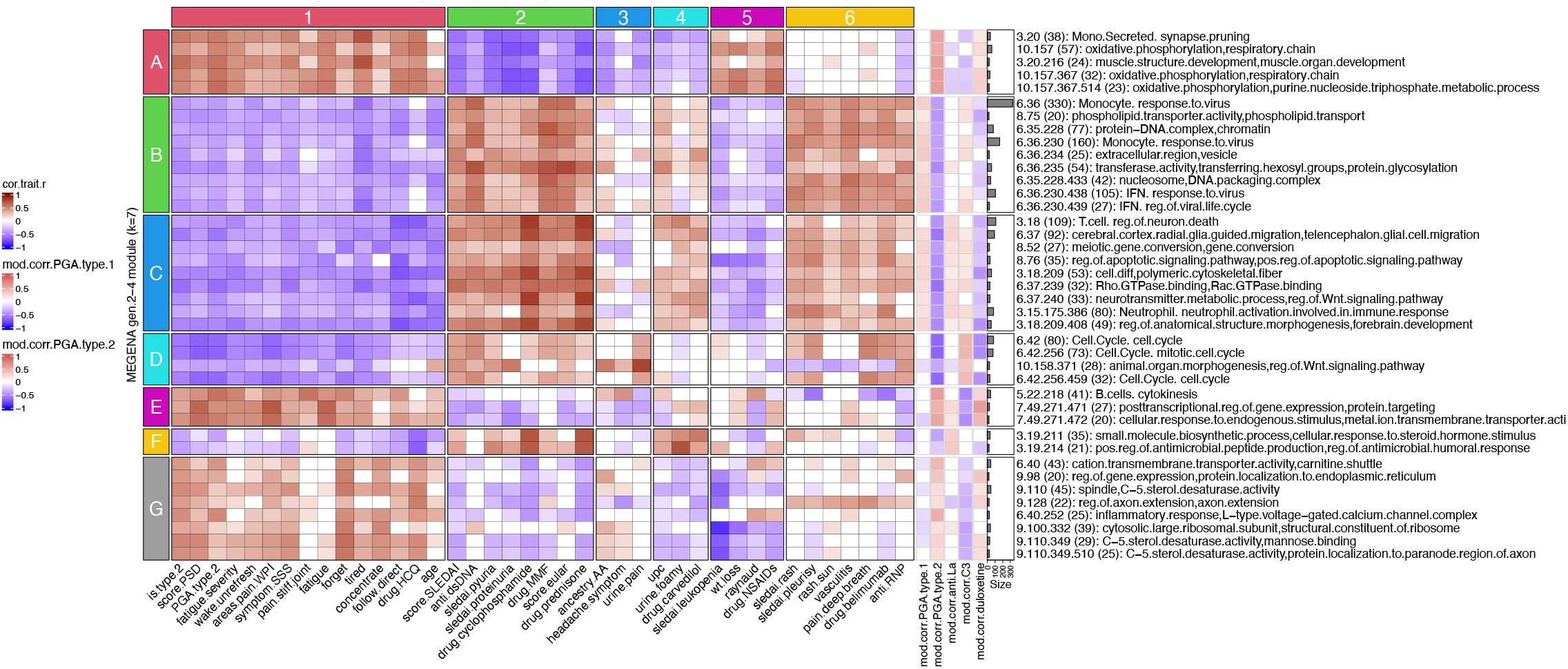
MEGENA module eigengene correlations to clinical & demographic features reveal specific gene modules associated with individual clinical characteristics of Type 1 and Type 2 SLE. Numerically encoded sample/patient traits were correlated to the first principal components (equivalent to the module eigengene ME) of all gen2 through gen4 MEGENA modules followed by identification of the top 40 significant (p<0.05) correlations to cohort (type 1 SLE vs type 2 SLE). The top 40 sample trait correlations were identified by descending rank order of absolute values of the summed correlations per each module row and are shown in the main heatmap portion of the figure. In the right portion of the figure, row annotations are shown for sample traits that were not included in the top 40 correlations, but are of clinical interest. These include ME correlations to PGA for type 1 SLE, PGA for type 2 SLE (seen in the central heatmap of the figure but repeated on the right side for ease of visual comparison), autoantibodies anti-La/SSB, low complement C3, and usage of duloxetine.

Because there was a numeric but not significant disparity in age between the groups (Type 1, 36.9+/−10.8 Type 2, 46.0 +/−8.7, p=0.07), we carried out two additional analyses to confirm that age was not contributing to the results. First, we eliminated the two youngest patients from the Type 1 group and the two oldest from the Type 2 group and repeated the analysis, resulting in a very similar separation of clinical features (**Supplementary Figure S5**).

Secondly, we used the entire group of patients and carried out the same analysis after covariant adjustment for age, again with similar results (**Supplementary Figure S6**). These results are all consistent with the conclusion that expression of co-expression modules is uniquely correlated with specific features of Type 1 and Type 2 SLE independent of age.

### Protein-protein Interaction (PPI) Analysis Identifies Biologic Function of Co-expression Modules

To provide insight into the biologic functions of genes within co-expression modules, we assessed genes within the top 40 MEGENA modules for PPIs using the STRING database (17). We found that 34 of the top 40 co-expression modules contained genes that were intraconnected by known PPIs, with 25 exhibiting 10-50% and 5 having > 50% PPI intraconnectedness (**Supplementary Table 4**). This finding confirms that the co-expression modules have captured known molecular pathways in an unsupervised manner. Type 1 SLE PPI intraconnected modules included cell cycle, T cells, regulation of neuronal death, extracellular region/vesicles, IFN and monocytes. Type 2 SLE PPI intraconnected modules included monocyte secretion, cation transport, axon extension, muscle structure development, and the inflammatory response/voltage gated calcium channel complexes. PPI connectedness is included as module row annotations in figures 3 and 4.

**Figure 3.**
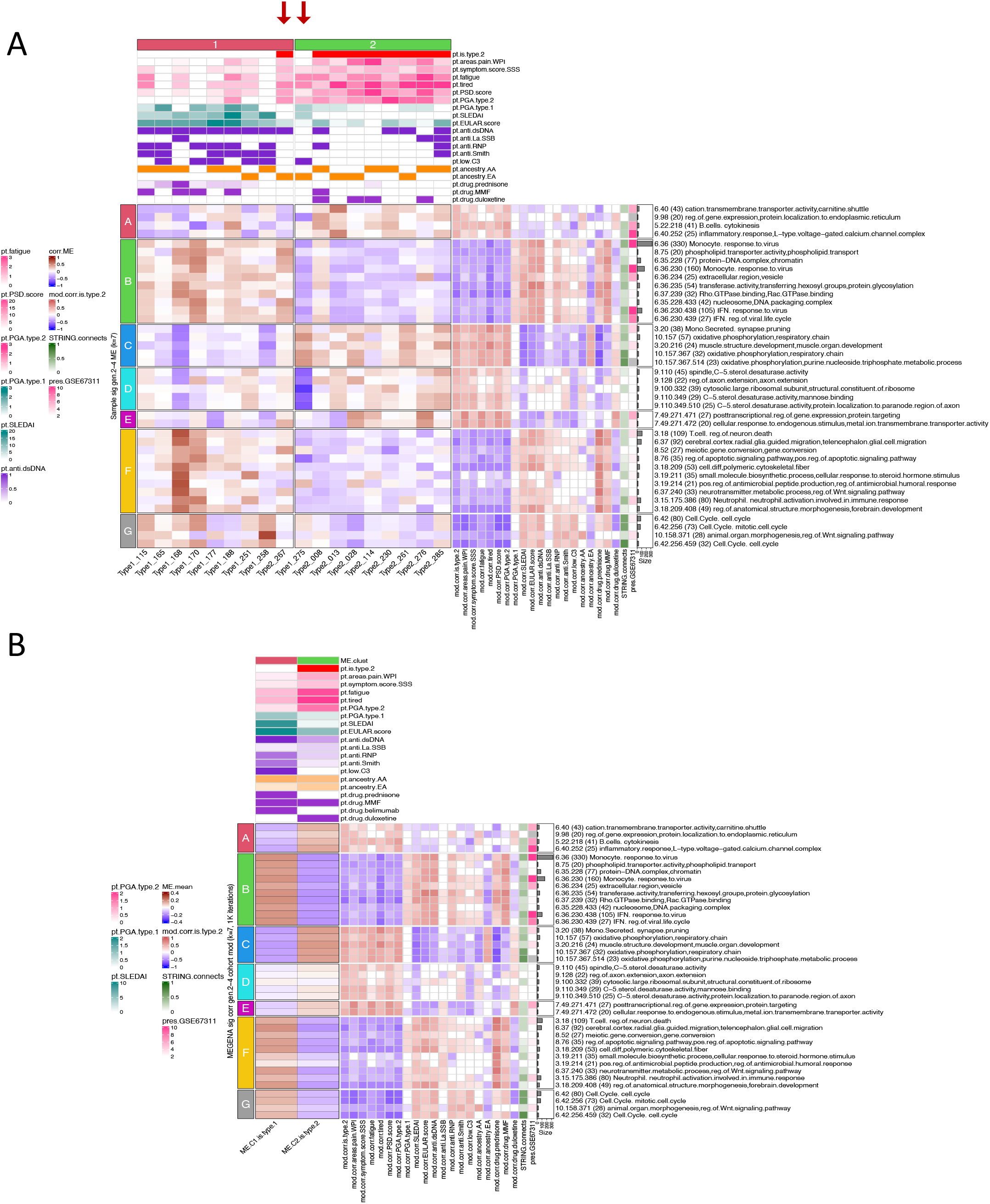
Clustering patient samples based on module eigengene (ME) separates type 1 SLE from type 2 SLE. The top 40 significant (p<0.05) type 1/2 SLE gen.2-4 cohort MEs were used to group patients using stable k-means clustering (k=2). Patient column annotations include patient type (type.1.SLE white, type.2.SLE red), areas of pain as measured by the widespread pain index (WPI), symptom severity score (SSS), PSD score, PGA for type 1 or type 2 SLE, SLEDAI score (with lab), ACR EULAR score, autoantibodies (anti-dsDNA, anti-LA/SSB, anti-RNP/SSA, anti-Smith), low C3 (binary), ancestral background (AA and EA), prednisone dosage, and usage of MMF or duloxetine (binary). Columns of sample traits were clustered using stable k-means clustering (k=2) with 1K iterations. Module rows were clustered in a similar manner on k=7 and include correlations to patient traits (−1 blue to +1 red), percentage of a given module’s genes participation in predicted protein-protein interactions per STRING analysis, and degree of module preservation in GSE67311 classic fibromyalgia (FM). Patients type1 275 and and type2 267 (red arrows) correspond to the same outliers identified during PCA analysis in supplementary figure S1 (**A**). Data from A was plotted as a mean of the patients in each cluster **(B)**. Column annotations are means of column annotations in (A) and row annotations are identical to row annotations in (A).

**Figure 4.**
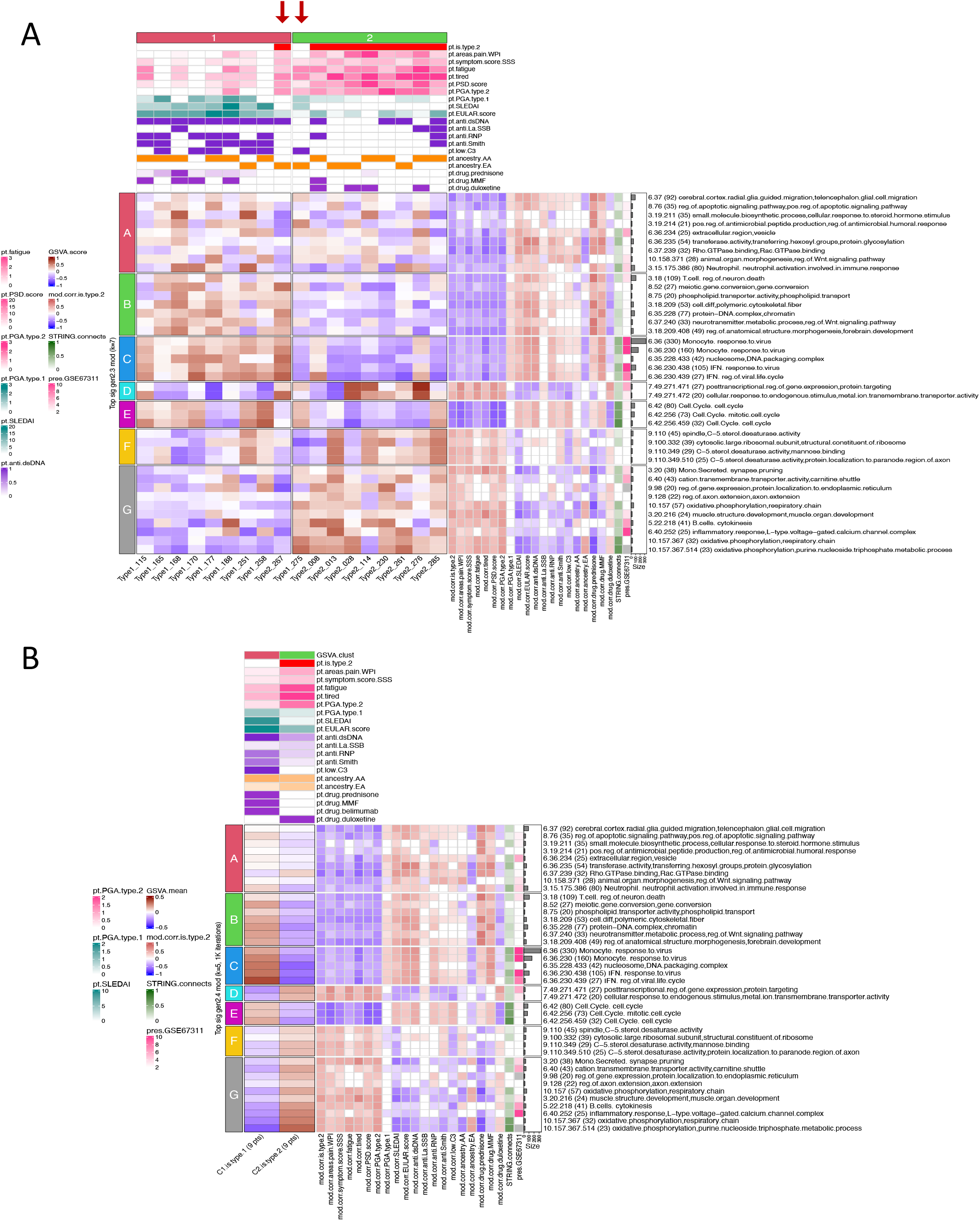
GSVA using MEGENA modules as input gene sets effectively separates Type 1 SLE from Type 2 SLE. Heatmaps indicate GSVA enrichment scores per patient for each module. Patient column annotations include patient type (type.1.SLE white, type.2.SLE red), areas of pain as measured by the widespread pain index (WPI), symptom severity score (SSS), PSD score, PGA for type 1 or type 2 SLE, SLEDAI score (with lab), ACR EULAR score, autoantibodies (anti-dsDNA, anti-LA/SSB, anti-RNP/SSA, anti-Smith), low C3 (binary), ancestral background (AA and EA), prednisone dosage, and usage of MMF or duloxetine (binary). Columns of sample traits were clustered using stable k-means clustering (k=2) with 1K iterations. Module rows were clustered in a similar manner on k=7 and include correlations to patient traits (−1 blue to +1 red), percentage of a given module’s genes participation in predicted protein-protein interactions per STRING analysis, and degree of module preservation in GSE67311 classic fibromyalgia (FM). Patients type1 275 and and type2 267 (red arrows) correspond to outliers identified during PCA analysis in supplementary figure S1 (**A**). Data from A was plotted as a mean of the patients in each cluster (**B**). Column annotations are means of column annotations in (A) and row annotations are identical to row annotations in (A).

### Gene Co-expression Modules Distinguish Type 1 and Type 2 SLE

Stable K-means clustering of co-expression module MEs was used to determine whether Type 1 and Type 2 SLE patient samples could be distinguished. Effective separation of Type 1 and Type 2 SLE patients was achieved, with only two outliers (Type1_275 and Type2_267) noted (**Figure 3**). Unique patterns of co-expression module MEs and Type 1 and Type 2 SLE, respectively, can clearly be seen. Moreover, unique and opposing co-expression module ME correlations with SLEDAI and PSD scores or PGA Type 2 were found. Notably, MEs of co-expression modules identifying the interferon signature and monocytes were highly positively correlated to SLEDAI and negatively correlated to PSD score. Conversely, the MEs of the axon extension, muscle structure development and B cell modules were negatively correlated to SLEDAI and positively to PSD score. Finally, patient ancestry also was correlated with specific co-expression module MEs. The detailed correlations between the coefficients of specific gene module expression and clinical traits are shown in supplemental figures S3 and S4 and confirm the largely mutually exclusive relationship between co-expression module expression and Type 1 or Type 2 features.

### Gene Co-expression Modules Distinguish Type 1 and Type 2 SLE using GSVA

To confirm this finding in an orthogonal manner, we used Gene Set Variation Analysis (GSVA) using co-expression modules as input gene sets followed by stable k-means clustering of GSVA scores. This approach also effectively distinguished Type 1 and Type 2 SLE patients (**Figure 4**).

### DEG Pairs Distinguish Type 1 and Type 2 SLE Samples

We employed Differential Gene Coexpression Analysis (DGCA) (18) as a complementary method to distinguish patients with active Type 1 or Type 2 SLE symptoms in greater detail. Here, DGCA was used to detect intermodular pairs of genes as a way to delineate potential differences between the molecular communication inherent in Type 1 and Type 2 SLE pathology. As seen in **Supplementary Figure S7** and **Supplementary Tables 5 & 6**, top unique intermodular connections distinguished Type 1 SLE from Type 2 SLE patients. Type 1 SLE patients were remarkable for neutrophil involvement/cell activation immune response and monocytes and Type 2 SLE patients largely for B cell interactions.

### Co-expression Module Preservation Between Type 1 and Type 2 SLE and FM Samples

Next, we sought to determine the relationship between the co-expression modules used to distinguish Type 1 and Type 2 SLE and those generated from a dataset of classic FM (GSE67311). MEGENA was employed to generate co-expression modules from the 70 FM patient samples in this dataset, and the MEs of the top 40 modules correlating to the seven clinical traits (bipolar disorder, BMI, CFS, FIQR, IBS, migraine, major depression) were visualized (**Supplementary Figure S8 A**). Module preservation was then carried out between the Type 1 and Type 2 SLE co-expression modules and those generated from GSE67311 FM samples. Using a composite z summary score (**Supplementary Figure S8 B**), 40 of the 157 Type 1 and Type 2 SLE modules were preserved (z score >2), 29 were moderately preserved (z score >5), and 21 were well preserved (z score >10) among the FM co-expression modules. Functional annotations of top preserved modules showed immune/inflammatory cells, including monocytes, T cells, neutrophils, functional activities, including IL-1, cytokines, MHC binding and IFN, and also glial cell migration and axon guidance (**Supplementary Table 7**). The degree of module preservation in GSE67311 patients is included as module row annotations in figures 3 and 4.

### GSVA Further Distinguishes Type 1 and Type 2 SLE Patients and Identifies a Subset of Fibromyalgia (FM)

We next assessed in greater detail the relationship between SLE gene expression abnormalities and those in FM. For this purpose, we used stable k-means clustering of GSVA scores to generate five distinct FM patient clusters (**Figure 5**). Notably, a subset of idiopathic FM patients (18/45, 40%) molecularly resembled Type 2 SLE patient signatures (patients within vertical clusters 1 and 3), and “fatigue” and “tired” Type 2 SLE modules were highly correlated to this patient subset. Co-expression modules included strong correlations in opposing directions to patients with Type 1 SLE vs Type 2 SLE symptoms. Type 1-like FM patients were notably and positively correlated to the horizontal module clusters A, C, and D that included monocytes, IFN, T cells, cell cycle, neutrophils, and neurotransmitter processes. Type 2-like FM patients were notably and positively correlated to the horizontal module clusters E, G, and H that included metabolic pathways, muscle structure development, B cells, and L-type voltage-gated calcium channel complexes.

**Figure 5.**
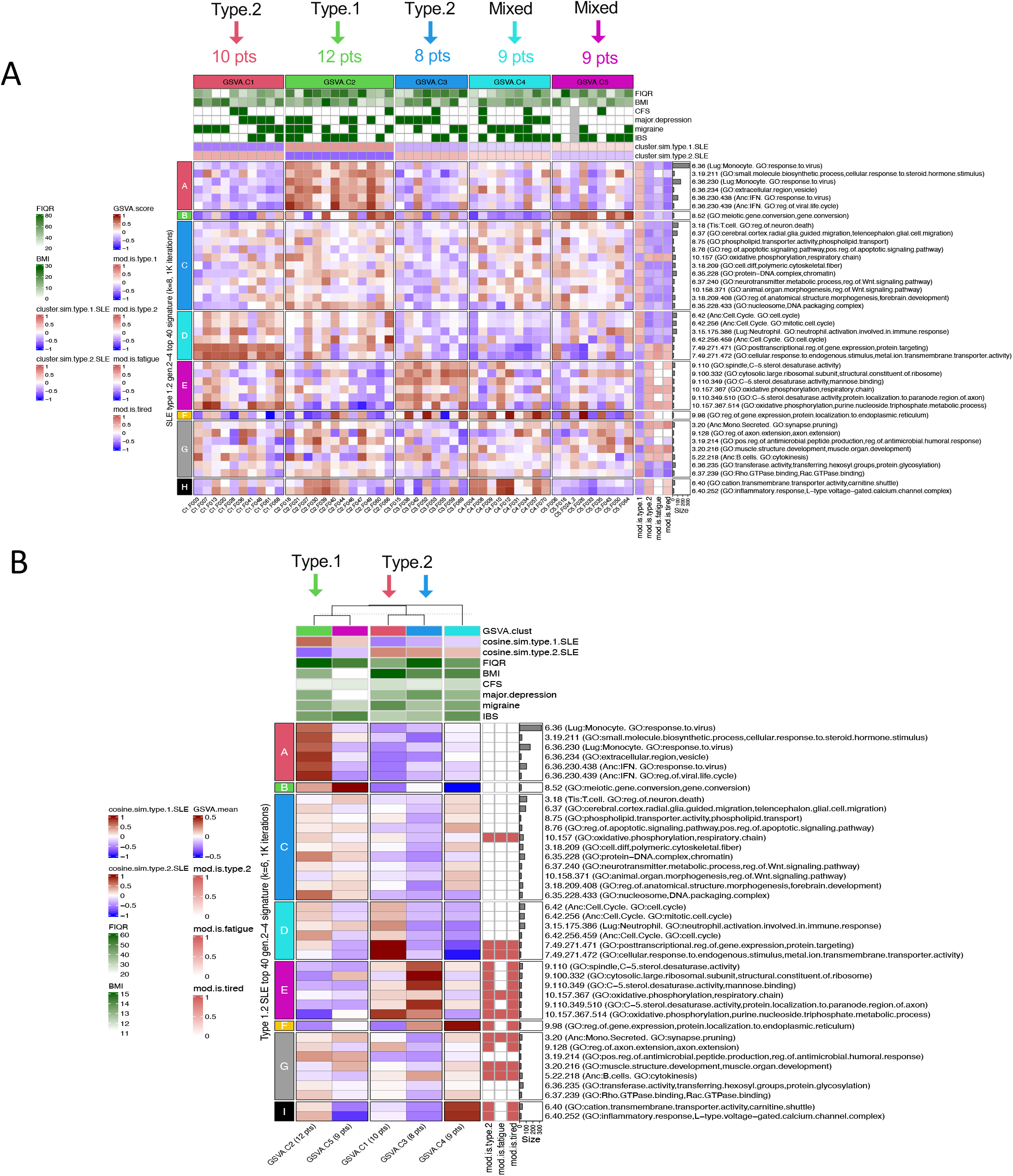
Type 1/2 SLE molecular signatures identify 18/45 (40%) of subjects with fibromyalgia (FM) from GSE67311 exhibiting enrichment of type 2 gene modules. The top5k rowVar genes from GSE67311 were analyzed by GSVA using the top 40 type 1/2 SLE cohort gen.2-4 modules as gene signatures. Column annotations include FIQR (Fibromyalgia Impact Questionnaire Score), BMI (body mass index), CFS (chronic fatigue syndrome), major depression (yes/no), migraine (yes/no), IBS (irritable bowel syndrome yes/no), and mean cluster cosine similarity to bona fide type 1 and type 2 sample results. FM patient cluster 2 (12 patients) is most similar (cosine sim > 0.3) to type 1 SLE signatures, and FM patient clusters 1 (10 patients) and 3 (8 patients) are most similar to type 2 SLE signatures. Clusters 4 and 5 had only weak similarity type 1 or type 2 SLE (sim < 0.3). Row annotations indicate modules that were significantly correlated to type 1 SLE or type 2 SLE, fatigue, and tired. Columns were stably clustered (1k iterations) into k=5 patient clusters and rows optimally clustered into k=8 groups of modules (**A**). GSVA enrichment score row means and sample traits were calculated for the five GSVA patient clusters (**B**).

### Type 1 and 2 SLE Modules Identify a Subset of Inactive SLE Patients

We next determined whether patients with the Type 2 SLE signature could be found in other datasets of patients (GSE45291 and GSE49454) with inactive SLE (SLEDAI<6). Stable k-means clustering based on GSVA scores using the Type 1 and Type 2 SLE co-expression clusters formed four distinct groups within each study. In GSE45291, most inactive SLE patients (151/244, 61.8%) were not identified by GSVA using Type 1 and Type 2 co-expression modules as gene sets. However, 49/244 (20.1%) inactive SLE patents were identified by enrichment of the Type 2 co-expression modules (**Figure 6)**. Notably, a similar number (44/244, 18%) were identified by enrichment of the Type 1 gene signature. Similar results were seen in inactive SLE patients in GSE49454 (**Supplementary Figure S9**). These results indicate that most patients with inactive SLE do not express either the Type 1 or Type 2 gene expression signature. However, small subsets express one or the other, suggesting that a small proportion of each may have the molecular profile of Type 1 and 2 SLE. A summary of the distribution of inactive SLE and FM patients showing molecular features of Type 1 and Type 2 SLE is shown in **Supplementary Figure S10**. Unfortunately, clinical features of Type 2 SLE are not available in these datasets.

**Figure 6.**
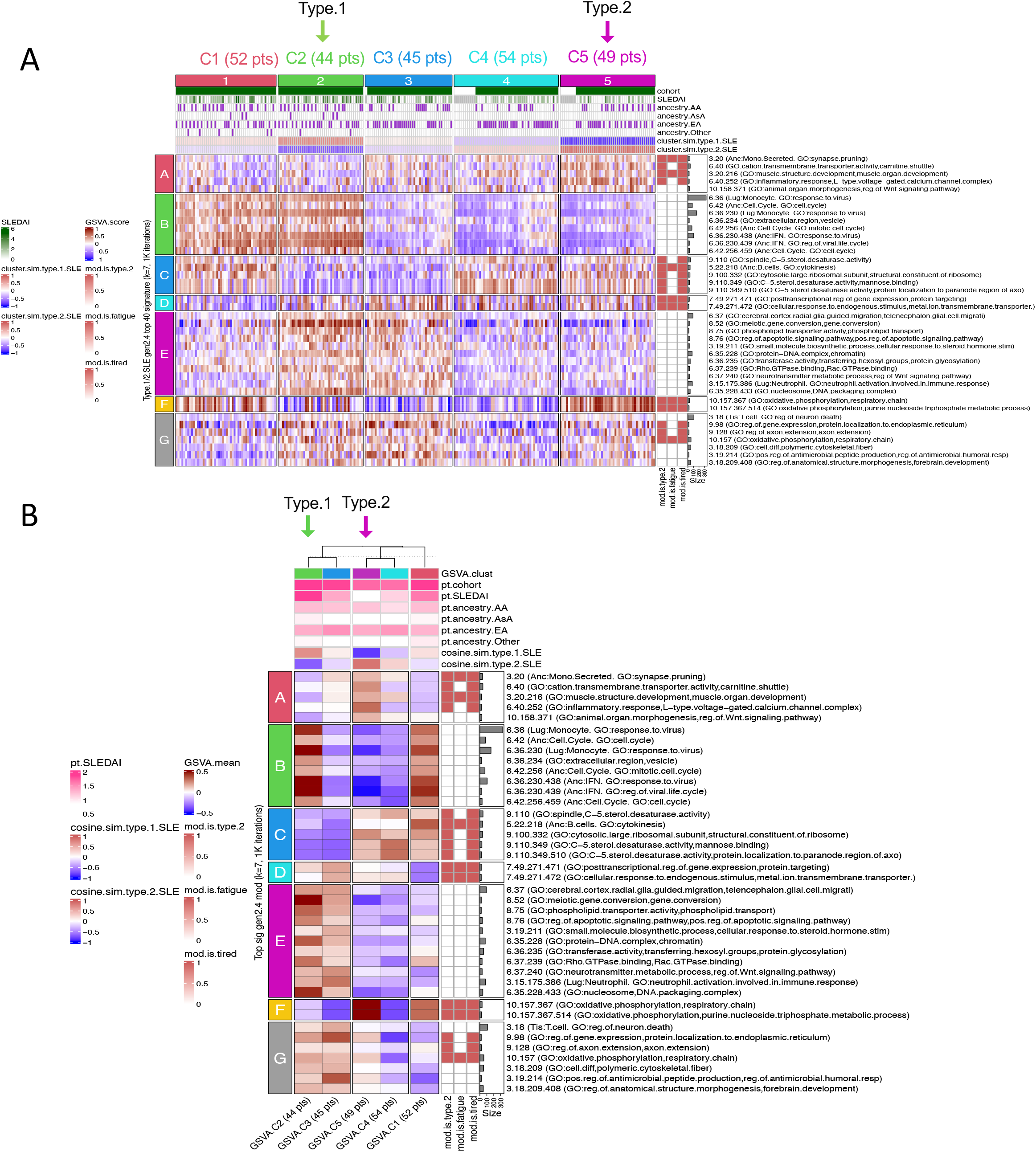
Type 1/2 SLE molecular signatures identify a small subset of 49/244 (20.0%) of subjects with inactive SLE (SLEDAI <6) from GSE45291 exhibiting enrichment of type 2 gene modules. The top5k rowVar genes from GSE45291 were analyzed by GSVA using the top 40 type 1/2 SLE cohort gen.2-4 modules as gene signatures. Column annotations include cohort (healthy or SLE), SLEDAI score and ancestral background (AA African ancestry, AsA Asian ancestry, EA European ancestry, and other), and mean cluster cosine similarity to bona fide type 1 and type 2 sample results. Inactive SLE patient cluster 2 (44 patients) is most similar to type 1 SLE signatures and inactive SLE patient cluster 5 (49 patients) is most similar to type 2 SLE signatures. Clusters 1, 3, and 4 had only weak similarities to type 1 or type 2 SLE (sim < 0.3). Row annotations indicate modules that were significantly correlated to type 2 SLE, fatigue, and tired. Columns were stably clustered (1k iterations) into k=5 patient clusters and rows optimally clustered into k=7 groups of modules **(A)**. Mean GSVA enrichment scores and sample traits were calculated for the five GSVA patient clusters **(B)**.

### SLE Subsets Identified by Type 2 SLE Gene Modules Have Severe Fatigue More Frequently

Finally, we sought to determine whether subsets of SLE patients identified by enrichment of Type 2 SLE modules have a greater frequency of severe fatigue. We employed GSE88884 (Illuminate 2) for this analysis even though this dataset set was limited to patients with active disease (SLEDAI of 6 or more) because fatigue and pain were measured, albeit using different metrics (Brief Fatigue Inventory and Brief Pain Inventory). As can be seen in **Figure 7**, using k-means clustering based on enrichment of the 40 SLE Type 1 and 2 co-expression modules and GSVA, GSE88884 samples were separated into 6 subsets, 2 with similarity to Type 2 SLE, 1 with similarity to Type 1 SLE, and 3 with mixed features. When these subsets were interrogated for the frequency of severe fatigue, the two Type 2-like subsets were significantly enriched for patients with severe fatigue along with one of the mixed subsets. Further analysis of this mixed subset indicated minimal or no enrichment of the horizontal module cluster G containing monocyte and interferon signatures. It is notable that all subsets contained significantly more patients with mild pain with no differences between the subsets.

**Figure 7.**
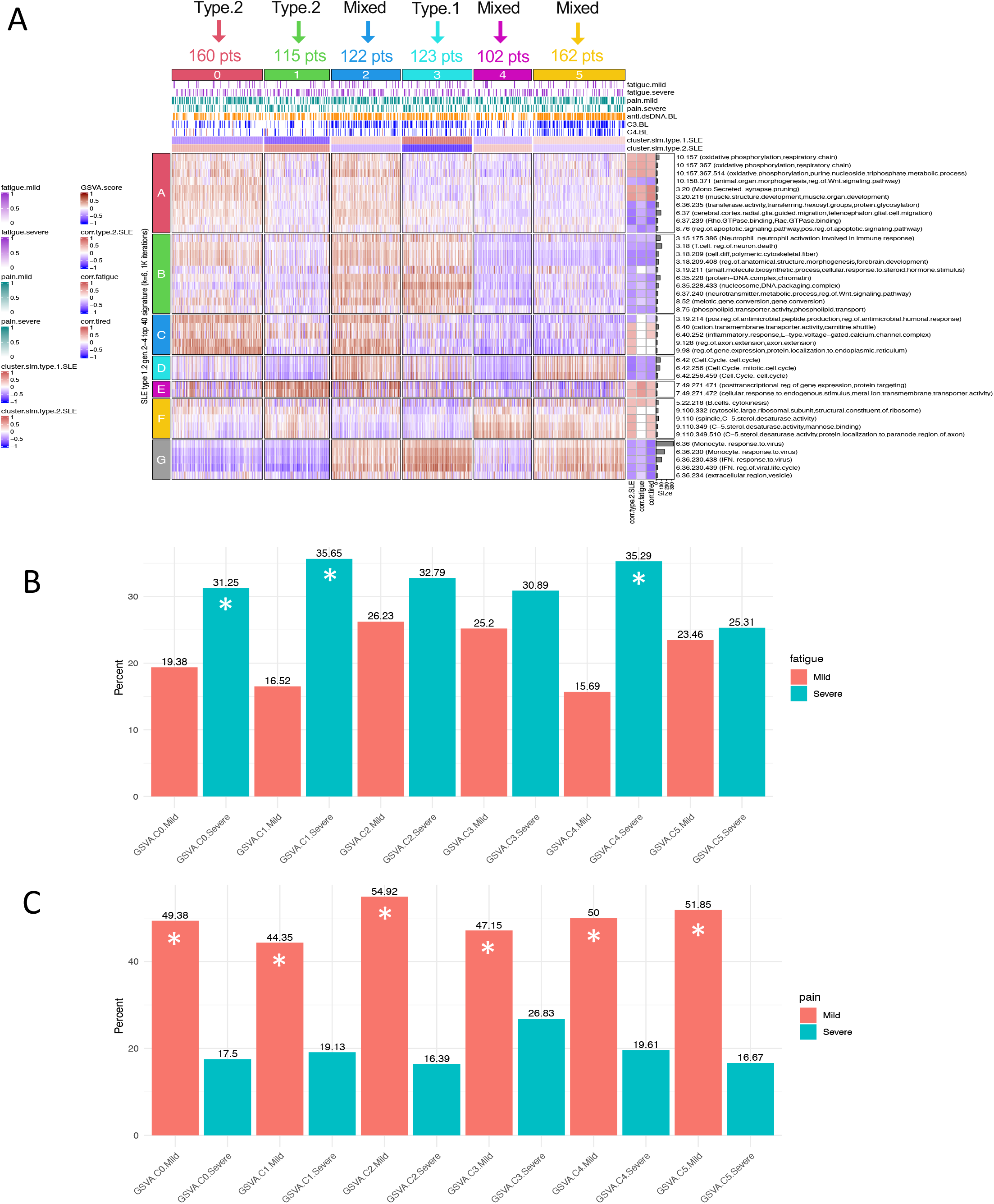
Analysis of patients with active SLE (GSE88884) identifies patient groups with severe fatigue. GSVA was carried out on GSE88884 (ILLUM-2) using the top 40 type 1/2 SLE cohort modules as signatures. Stable k–means clustering of GSVA enrichment scores formed 6 patient clusters and 6 module clusters. Column annotations include mild or severe fatigue (mild 1-3, severe 8-10) using the Brief Fatigue Inventory, mild or severe pain scored using the Brief Pain Inventory (mild 1-4, severe 7-10), anti-dsDNA, C3 and C4 at baseline (low −1, normal 0, high +1), and mean cluster cosine similarity to the Type 1 SLE & Type 2 SLE patient clusters. ILLUM-2 patient cluster 3 was most similar by cosine similarity to type 1 SLE signatures, and clusters 0 & 1 were most similar to type 2 SLE signatures. Clusters 2, 4, and 5 were mixed (type.2.SLE cosine similarities −0.34, +0.36, and −0.23, respectively). Row annotations indicate modules that were significantly correlated to type 1/2 SLE, fatigue, and tired (**A**). Proportion test analysis significantly (p<0.05) identifies ILLUM-2 patient groups with fatigue by the Brief Fatigue Inventory (mild 1-3, severe 8-10) (**B**) and those with pain scored using the Brief Pain Inventory (mild 1-4, severe 7-10) (**C**). Patient clusters marked as (*) exhibit a significant difference between the frequency of severe and mild fatigue or pain, respectively.

## DISCUSSION

In this pilot study using a bookend approach, we tested the hypothesis that patients with SLE with high levels of Type 1 or Type 2 symptomatology can be distinguished on the basis of transcriptomic analysis of peripheral blood cells. While the number of patients in this study was limited, the data nevertheless support five important conclusions concerning Type 1 and Type 2 SLE activity. First, co-expression gene modules derived from Type 1 and 2 SLE patients highly correlate with specific features of Type 1 and 2 SLE. Secondly, patients with active Type 1 or Type 2 SLE have quite distinct gene expression profiles, with perturbations of specific molecular pathways. Thirdly, the Type 1 and Type 2 SLE-related gene expression profiles can identify unique subsets of FM patients. Fourthly, the gene expression profiles of Type 2 SLE can be detected in unrelated datasets comprised of patients with inactive SLE. Finally, the Type 2 SLE gene co-expression modules identify subsets of patients with active SLE with a greater frequency of severe fatigue.

Previous studies of peripheral blood cells have primarily addressed the relationship of changes in gene expression to inflammatory disease activity as measured by instruments such as the SLEDAI (19). These studies have thus focused largely on Type 1 disease. This raises the question of whether the differences in gene expression profiles merely are indicative of differences in disease activity. A number of studies have assessed gene expression changes related to changes in disease activity measured by SLEDAI. Although changes have been identified in different studies (20), no consensus pattern of gene expression has been determined (21) Moreover, in this study, the Type 2 gene expression profile was seen in only a small fraction of inactive patients in two datasets and also in a subset of SLE patients with active disease. Therefore, it is unlikely that the Type 2 gene expression profile merely reflects changes in SLEDAI score. In this regard, association of the interferon gene signature with Type 1 SLE is notable. In general, the interferon signature is associated with the diagnosis of SLE, but may not change significantly over time in longitudinal studies of adult patients with disease activity in individual pediatric patients (22–24). Of note, recent studies have revealed a significant association between the interferon signature and the presence of specific autoantibodies, especially those to RNA binding nuclear proteins, including anti-RNP, anti-Sm and anti-SSA (25). Notably, administration of Type 1 interferon as a therapeutic can cause symptoms consistent with Type 2 SLE activity, including fatigue and achiness (26). In the current study, an association was found between the interferon gene signature and Type 1 but not Type 2 SLE activity. These results clearly establish an association between the interferon signature and Type 1 SLE, consistent with the role of both interferon and autoantibodies in the inflammatory features of SLE, similar to results reported here (27).

Beyond the interferon gene signature, expression of other specific gene modules was shown to be useful in distinguishing Type 1 and Type 2 SLE activity. These findings were validated using a number of orthogonal analytic techniques, including module eigengene correlations, GSVA enrichment scores, and analysis of DGCA intermodular pairings. Unique Type 1 SLE gene module enrichments included monocytes, neutrophils, T cells, interferon, IL-1, TNF, cell cycle and Wnt signaling, all characteristic of the inflammatory nature of this form of SLE. DGCA more specifically implicated Type 1 SLE interactions between monocytes and neutrophils and a host of other neutrophil interactions, notably including IL-1 and IFN. DGCA also showed that cell cycle was paired with the generation of superoxide and hydrogen peroxide as part of the neutrophil innate immune response, steroid precursor generation for manufacture of many molecules including immune signals, and T cell and Fc receptor activity. These features are all typical of the inflammatory nature of Type 1 SLE symptoms as previously reported for active SLE in general (1).

In contrast to findings with Type 1 SLE, expression of a number of other gene modules characterized active Type 2 SLE symptoms. We found a number of neural features that distinguished Type 1 and Type 2 SLE activity. Unique Type 1 SLE module enrichments included those annotated as cerebral cortex microglial cell migration and neurotransmitter metabolism. DGCA more specifically suggested Type 1 SLE intermodular connections between neutrophils and neurotransmitter metabolism, postsynaptic endosomes, and nervous system development. It was initially surprising in this study of peripheral blood that one module was annotated as microglia rather than monocytes/macrophages. Although these cell types share no common progenitor, they are both members of the mononuclear phagocyte system and share functional features which could lead to overlaps in cell type annotations. Additional studies will be necessary to determine whether enrichment of this module reflects microglial or general monocyte/macrophage enrichment in Type 1 SLE, but this enrichment is consistent with previous studies on the contribution of mononuclear phagocyte activity to inflammatory features of SLE (28–30).

It is also of interest that Type 1 SLE activity was associated with a neutrophil signature. Previous studies have clearly delineated a role of neutrophil subpopulations in active SLE (31,32) and, notably, in this study, this association was only found in patients with active Type 1 and not Type 2 SLE. In addition, steroid usage was positively correlated to neutrophils, monocytes, IL-1, and the Fc-receptor in Type 1, but these features were all negatively correlated to Type 2 SLE. These finding implies that neutrophils may contribute to the features of Type 1 but not Type 2 SLE, although steroid administration is a possible contributor (21,22,33)

Type 2 SLE was also notable for neuromuscular and metabolism enrichments, sufficiently distinct to be detected in peripheral blood. These findings include muscle structure development, oxidative phosphorylation, cation transport, the carnitine shuttle (concentrated in skeletal and cardiac muscle), and L-type voltage gated calcium channel complexes (which are associated with skeletal, smooth, and cardiac muscle). Mitochondrial dysfunction and homeostatic imbalance have been investigated in FM as potentially modulating neuropathic pain through links with energy metabolism (34) including mitochondrial abnormalities in carnitine fatty acid metabolism (35). It has been suggested that there is a connection between reactive oxygen species (ROS) and neuropathic pain and that mitochondria could be a therapeutic target in FM and may also be involved in sensitivity to painful stimuli in Type 2 SLE (36,37).

Besides identifying gene expression modules that discriminate Type 1 from Type 2 SLE, we identified patient clusters derived from two studies of inactive SLE patients that shared some transcriptional patterns with those we found with Type 2 SLE. Only a small fraction of inactive SLE patients were enriched for the Type 2 gene signature (20.1-34.6%). Because we did not have information on Type 2 symptoms in these patients, we went on to analyze patients from a clinical trial (GSE88884, Illuminate 2) because fatigue and pain were recorded, even though all of these patients manifested active disease (SLEDAI >=6). It is notable that an increased frequency of severe fatigue was found in the subsets with Type 2 gene expression features and even in a subset with mixed molecular features but diminished Type 1 monocyte and interferon gene expression. It was surprising that no difference in the frequency of severe pain was noted in the subsets, but this could relate to differences in the information collected by the WPI versus the Brief Pain Inventory.

Our study is the first attempt to assess differences in gene expression in patients who have been selected to have primarily Type 1 SLE or Type 2 SLE at the time of analysis, a so-called bookend approach. All patients with current Type 2 SLE activity have had active Type 1 SLE in the past, as Type 1 activity is required to meet criteria for SLE (5,6). It is, therefore, interesting to speculate that Type 1 and Type 2 symptoms may vary in individual SLE patients and gene expression profiling may be useful to delineate or possibly even predict the transition. It is also possible that Type 1 and 2 symptoms may co-exist in some patients as fatigue, for example, is present in as many as 90% of all SLE patients, and that gene expression profiling might be useful in dissecting the molecular endotype of each set of manifestations. The preliminary analysis of patients with active SLE supports this conclusion.

Our study also indicates a relationship between transcriptional patterns in Type 2 SLE and a subset of FM patients, including enrichments of B cells, plasma cells, and IgG chains as identified using DGCA. Since many factors can lead to central sensitization, a key postulated mechanism for FM, it is not surprising that there is heterogeneity in the transcriptional profiles. The observation of common features in a subset of FM is, therefore, notable and suggests that despite diversity of causative factors for central sensitization, common transcriptional changes can occur whether FM occurs by itself or in the context of an inflammatory disease.

It is also of interest that a second subset of FM had a gene expression profile similar to that of Type 1 SLE. Notably, this subset had additional gene expression features of inflammation, including enrichments of monocytes, inhibitory macrophages, neutrophils, as well as interferon, TNF, and IL-1 pathways. Unfortunately, detailed clinical evaluations of these patients are not available to determine whether they did indeed have underlying inflammatory disease. Despite this uncertainty, the data suggest that gene expression profiling can distinguish subsets of FM, two of which are molecularly similar to Type 2 SLE, and a second with more inflammatory features typical of Type 1 SLE.

As a pilot study, the current study has limitations. The number of patients is relatively small. Moreover, we did not have detailed clinical information about subjects with FM or inactive SLE. Finally, we did not have the opportunity to follow patients longitudinally to determine whether molecular features track with or even precede clinical features of Type 1 and 2 SLE. Despite this, the results are provocative and merit confirmation in larger datasets.

In summary, our study utilized a number of orthogonal bioinformatics approaches to distinguish Type 1 from Type 2 SLE based on unique transcriptional patterns. Additionally, we identified a subset of Type 2 SLE-like patients in datasets of FM and inactive SLE, suggesting molecular similarities of these entities. Moreover, we could identify a subset of patients with active SLE who expressed the Type 2 gene expression profile and exhibited an increased frequency of severe fatigue. Finally, we found that a subset of FM patients showed molecular features of Type 1 SLE with upregulation of many inflammatory genes; these finding suggest the possibility of inflammatory components in some patients with idiopathic FM.

## Data Availability

All data produced in the present study are available upon reasonable request to the authors. Raw data files have been deposited in NCBI accession PRJNA858861.

https://www.ncbi.nlm.nih.gov/bioproject/?term=PRJNA858861

## Notes

**Financial disclaimer:** We hereby declare no financial support or other benefits from commercial sources for the work reported within this manuscript, or any other financial interests that any of the authors may have, which could create a potential conflict of interest or the appearance of a conflict of interest with regard to this work.

### Competing Interest Statement

The authors have declared no competing interest.

### Funding Statement

This study did not receive any funding

### Author Declarations

Duke University Health IRB Pro00008875 Duke University IRB Pro00094645

