## Supplementary Figure S1 for "The Molecular Endotypes of Type 1 and Type 2 SLE"

A

| Duke Patient Characteristics | | | | |
|---|---|---|---|---|
| Type.1 patients | 9 | | | |
| Type.2 patients | 9 | | | |
| Patient genders | 17 female, 1 male | | | |
| No. ancestral backgrounds | 2 (11 African ancestry: AA, 6 European ancestry: EA, 1 Hispanic ancestry: HA) | | | |
| Total no. sample attributes | 105 (see supplementary table) | | | |
| PC variance explained | PC1 | PC2 | PC3 | PC4 |
| | 20.9% | 16.9% | 15.3% | 13.2% |

B

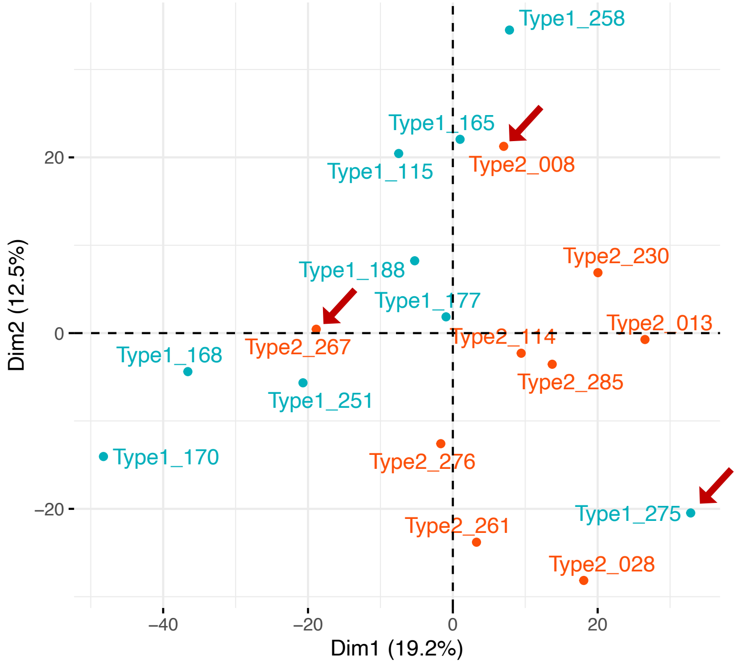


C

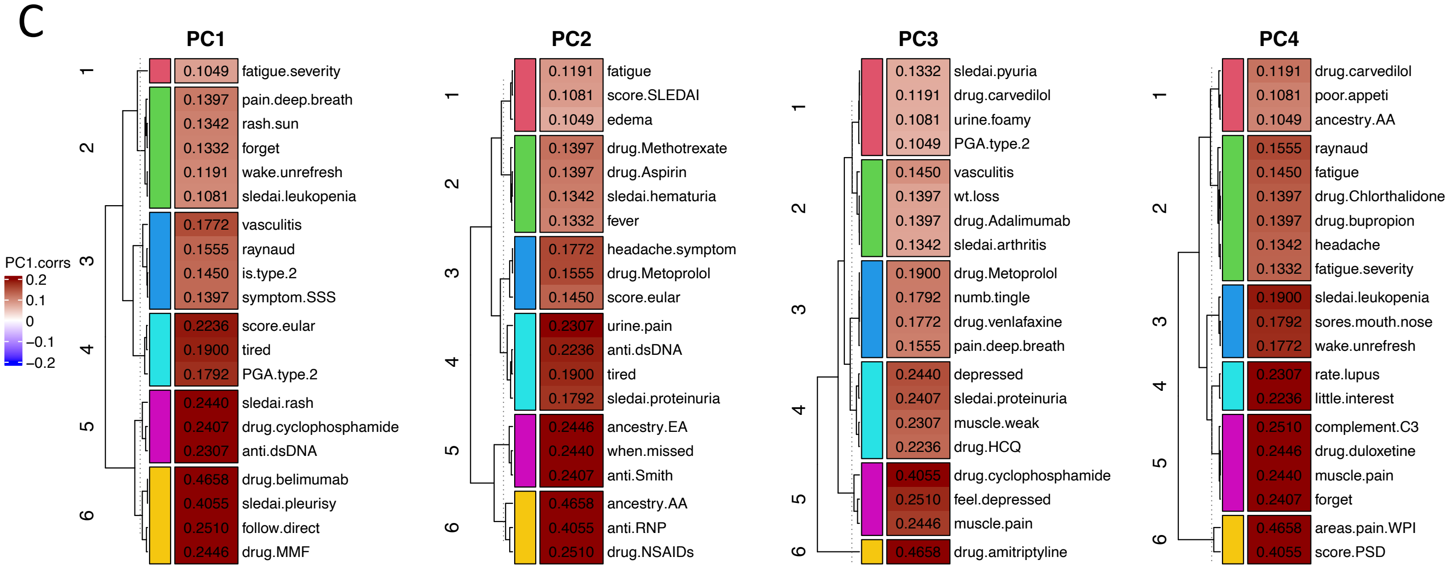


**Supplementary figure S1.** Patient heterogeneity and gene expression dimensionality reduction using principal component (PC) analysis of variance. Table of study participants and explained variance of first 4 PCs totaling to 66.3% (A). Biplot of PC2:PC1 of top 5k rowVar gene normalized expressions. Sample points colored by patient type. Red arrows indicate PCA outliers (B). Top 20 patient clinical/molecular attributes correlating to each of the top 4 PCs. Cells indicate r^2 correlation values and range from -0.2 to +0.2. Trait rows clustered by k=6 Euclidean distances (C).
