## Supplementary Figure S2 for "The Molecular Endotypes of Type 1 and Type 2 SLE"

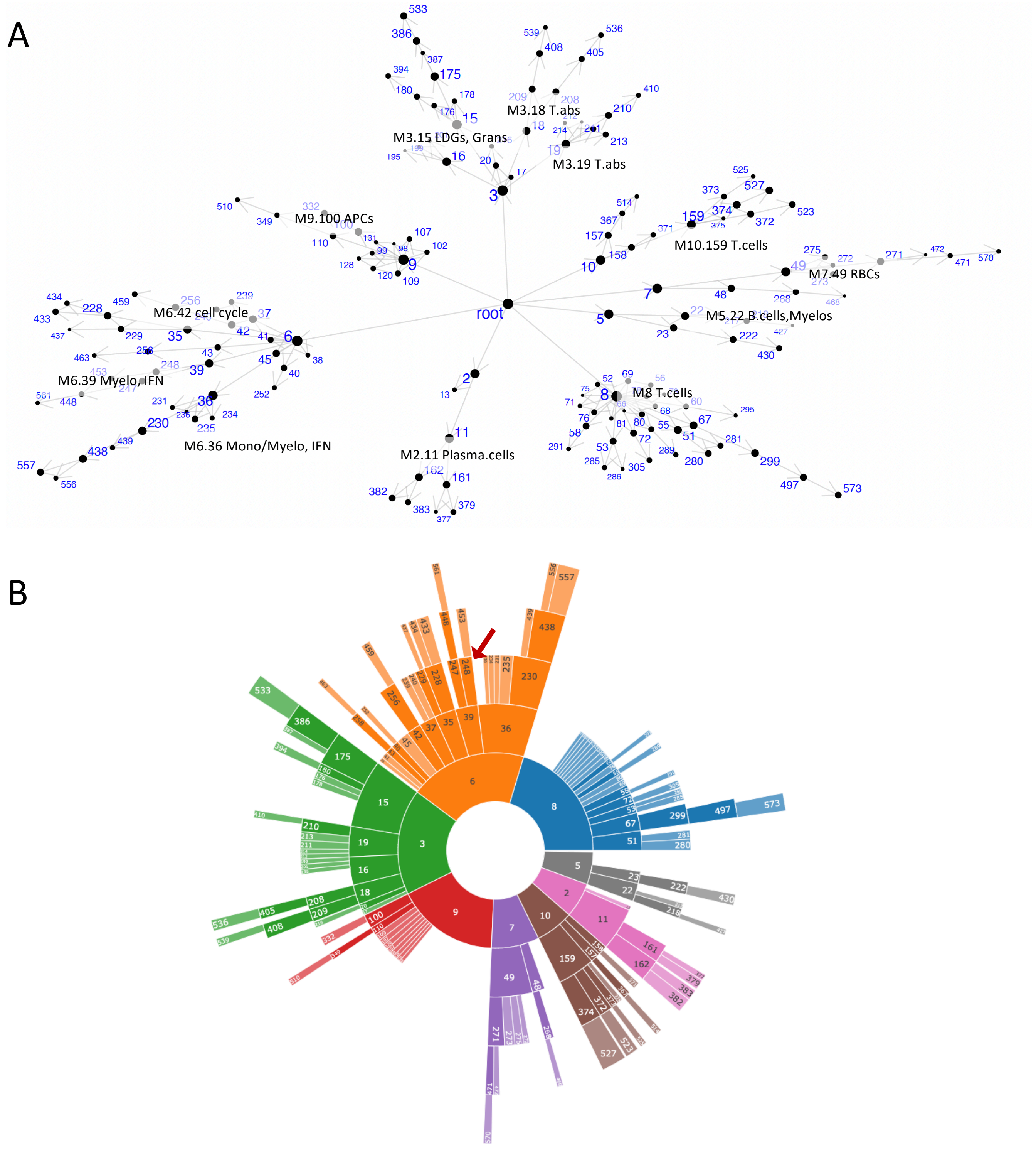


**Supplemental figure S2.** MEGENA coexpression module hierarchy visualizations. The top 5,000 row variance genes mapping to known proteins used to generate modules containing genes that were significantly intracorrelated by gene expression. The modules were iteratively clustered from the initial founder root modules and the lineages depicted as module nodes connected by straight interconnecting lines indicating module descendance (A). Modules were functionally annotated by statistically overlapping their gene symbols with lists of unique cell type or biological pathway gene markers. Functional designation required a minimum overlap of >=3 gene symbols and the overlap  Fisher's exact test ($p < 0.2$) to discard overlaps that occurred because of random chance. The architecture of module lineage is shown in (B) with modules pseudocolored by descendance to depict their linear relationships.
