## Supplementary Figure S3 for "The Molecular Endotypes of Type 1 and Type 2 SLE"

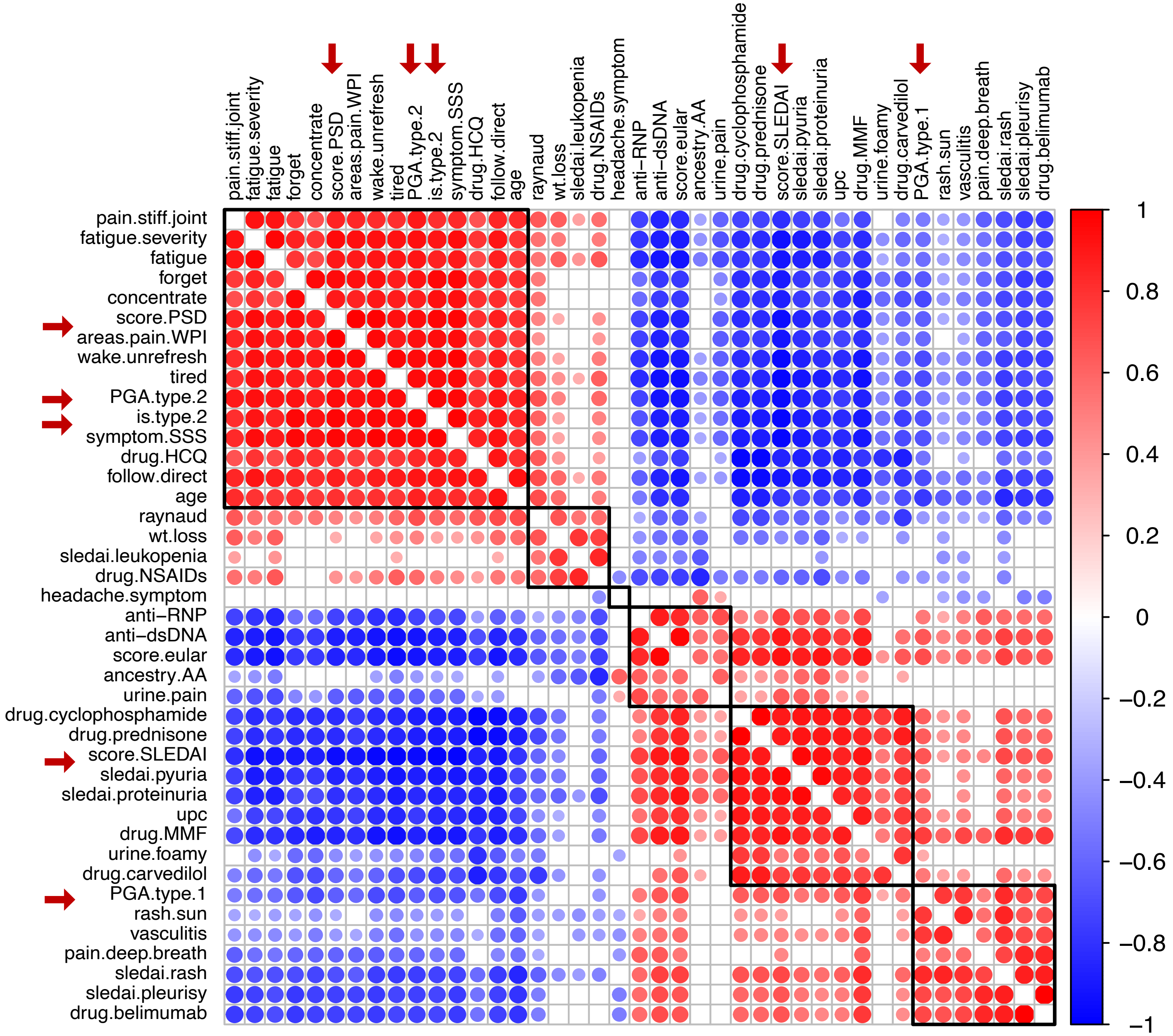


**Supplementary figure S3.** Plot of significant ($p<0.05$) correlations of expression of the top 40 cohort module MEs to clinical and molecular attributes. Red arrows indicate the clinical scores PSD, PGA.type.1, PGA.type2, SLEDAI, and is.type.2 cohort. Red indicates a positive correlation and blue indicates a negative one.
