## Supplementary Figure S4 for "The Molecular Endotypes of Type 1 and Type 2 SLE"

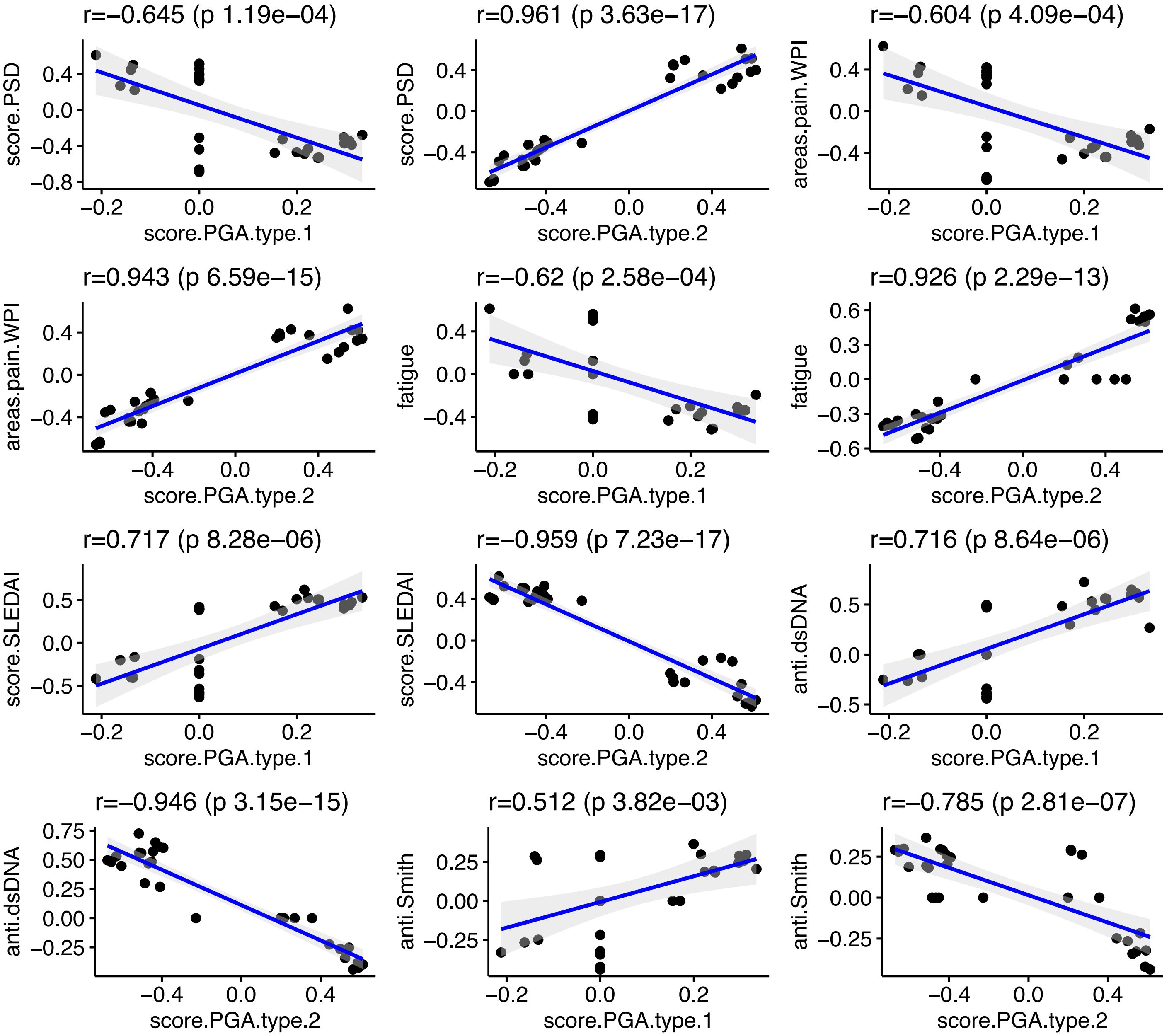


**Supplementary figure S4.** Correlations of the top 40 genes expression MEs with specific clinical features. The MEs of the top 40 gene expression modules were correlated with various clinical features and the correlation coefficients of the associations plotted on a two-dimensional matrix.
