## Supplementary Figure S5 for "The Molecular Endotypes of Type 1 and Type 2 SLE"

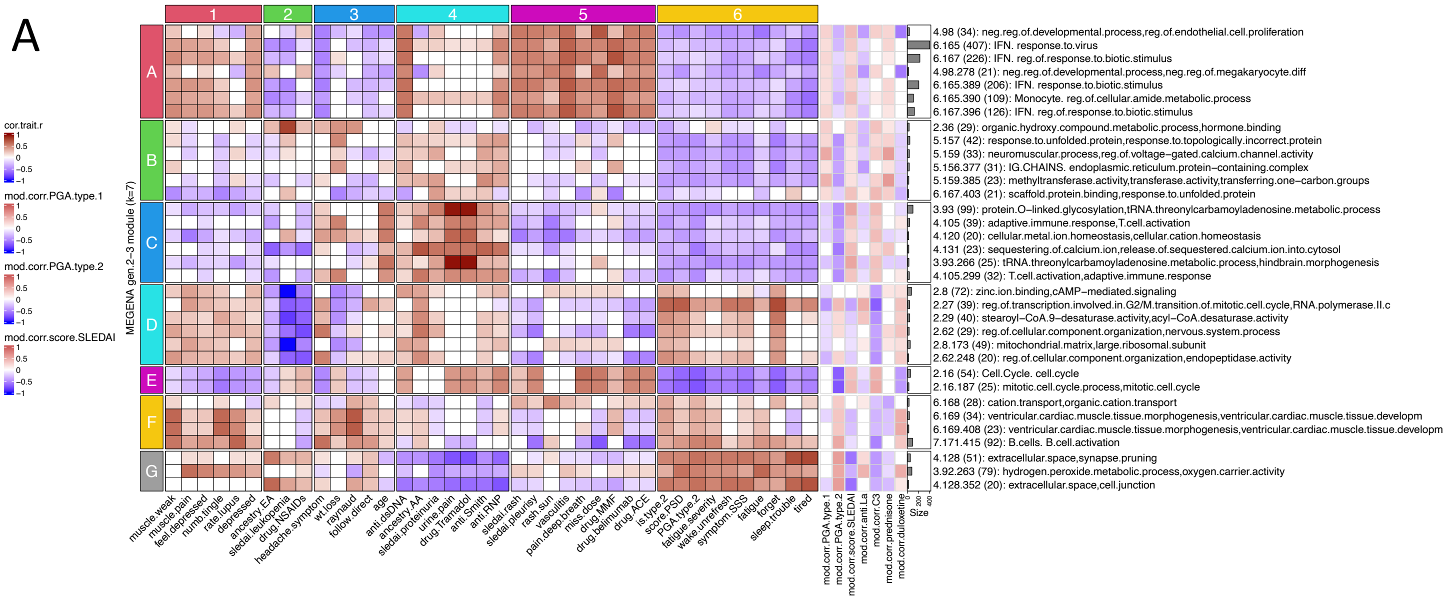


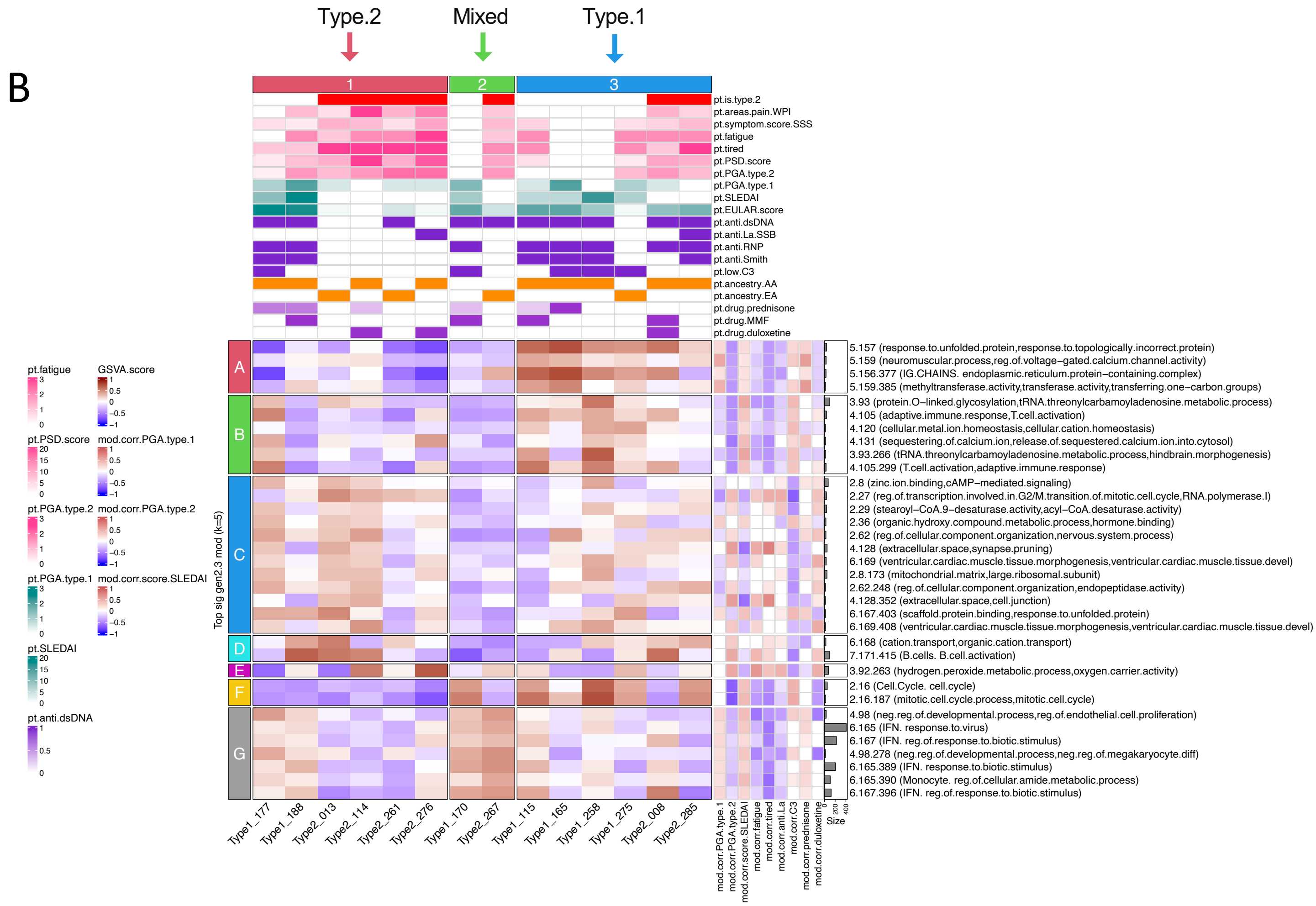


**Supplementary figure S5.** Removal of patient age outliers retains segregation of type 1 SLE from type 2 SLE clinical and molecular characteristics. Type 1 SLE patients 168 and 251, and type 2 SLE patients 028 and 230 were removed from the data set leaving 7 patients from each cohort with balanced age distributions. Gene expression of the 14 patients were submitted to MEGENA analysis forming new modules and the top 40 type 2 SLE ME module correlations visualized as a heat map (**A**). GSVA was performed on the top5k rowVar gene expressions using the top 40 age-balanced modules as signatures, forming distinct bookend groups of type 1 SLE patients (patient cluster 3), type 2 SLE (patient cluster 1), and patients with mixed molecular characteristics (patient cluster 2). Modules within the IFN module cluster G and Ig chain cluster A had overall positive correlations to type 1 SLE. Modules within the T cell cluster B more strongly negatively correlated to type 2 SLE. “Fatigue” and “tired” patient characteristics had positive correlations to type 2 SLE B cell cluster and erythrocyte modules. Patient columns were annotated with clinical and lab assay features identical to primary figures 3 and 4 (**B**).
