## Supplementary Figure S6 for "The Molecular Endotypes of Type 1 and Type 2 SLE"

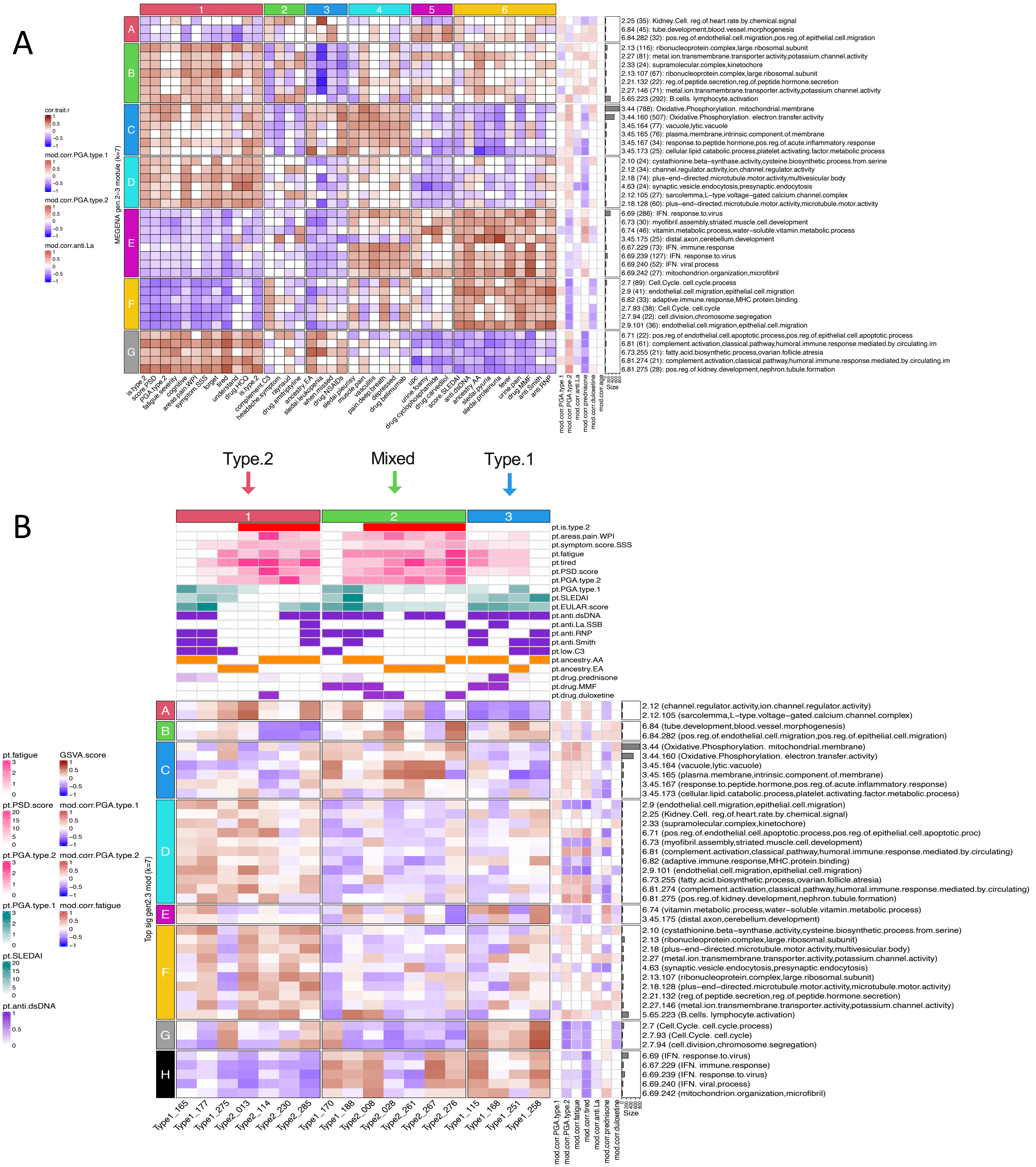


**Supplementary figure S6.** Age-adjusted gene expression retains segregation of type 1 SLE from type 2 SLE clinical and molecular characteristics. The top5k rowVar gene expressions of the original 18 patients were adjusted using linear modeling with age as a covariant. These were submitted to MEGENA analysis forming new modules and the top 40 type 2 SLE ME module correlations visualized as a heat map (**A**). GSVA was performed on the top5k rowVar gene expressions using the top 40 age-adjusted modules as signatures, forming groups of type 1 SLE patients (patient cluster 3), type 2 SLE (patient cluster 1), and patients with mixed molecular characteristics (patient cluster 2). "Fatigue" and "tired" patient characteristics had overall positive correlations to the type 2 SLE ox-phos, B cell, and complement pathway modules. "Tired" specifically and overall positively correlated to the type 2 SLE L-type voltage-gated calcium channel module, and both "fatigue" and "tired" were both overall highly negatively correlated to type 1 SLE IFN modules and cell cycle. Patient columns were annotated with clinical and lab assay features identical to primary figures 3 and 4 (**B**).
