## Supplementary Figure S7 for "The Molecular Endotypes of Type 1 and Type 2 SLE"

A

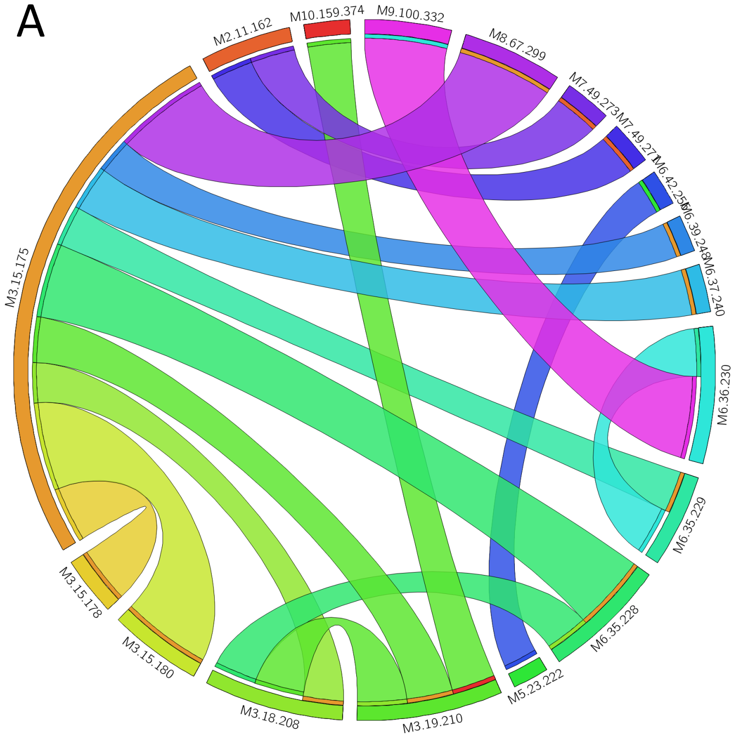


Top unique Type 1 SLE DGCA module pair members

| *lineage* | *enrichment* |
|---|---|
| 10.159.374 | GO:system.process,protein.heterodimerization.activity |
| 2.11.162 | Lug:Plasma-Cell. GO:response.to.unfolded.protein |
| **3.15.175** | **Lug:Neutrophil. GO:cell.activation.involved.in.immune.response** |
| 3.15.178 | GO:odontogenesis,trans-Golgi.network |
| 3.15.180 | GO:response.to.nitrogen.compound,reg.of.hormone.secretion |
| 3.18.208 | GO:postsynaptic.endosome,galactoside.binding |
| 3.19.210 | GO:GTPase.regulator.activity,interleukin-18-mediated.signaling.pathway |
| 5.23.222 | Anc:B.cells. GO:extracellular.matrix.organization |
| 6.35.228 | GO:protein-DNA.complex,chromatin |
| 6.35.229 | GO:nervous.system.development,RAGE.receptor.binding |
| 6.36.230 | Lug:Monocyte. GO:response.to.virus |
| 6.37.240 | GO:neurotransmitter.metabolic.process,reg.of.Wnt.signaling.pathway |
| 6.39.248 | Anc:IFN. GO:nucleic.acid.metabolic.process |
| 6.42.256 | Anc:Cell.Cycle. GO:mitotic.cell.cycle |
| 7.49.271 | GO:stem.cell.population.maintenance,maintenance.of.cell.number |
| 7.49.273 | GO:neg.reg.of.extrinsic.apoptotic.signaling.pathway,porphyrin |
| 8.67.299 | Tis:Monocyte/Myeloid.Cell. GO:pattern.recognition.receptor.signaling.pathway |
| 9.100.332 | GO:cytosolic.large.ribosomal.subunit,structural.constituent.of.ribosome |

B

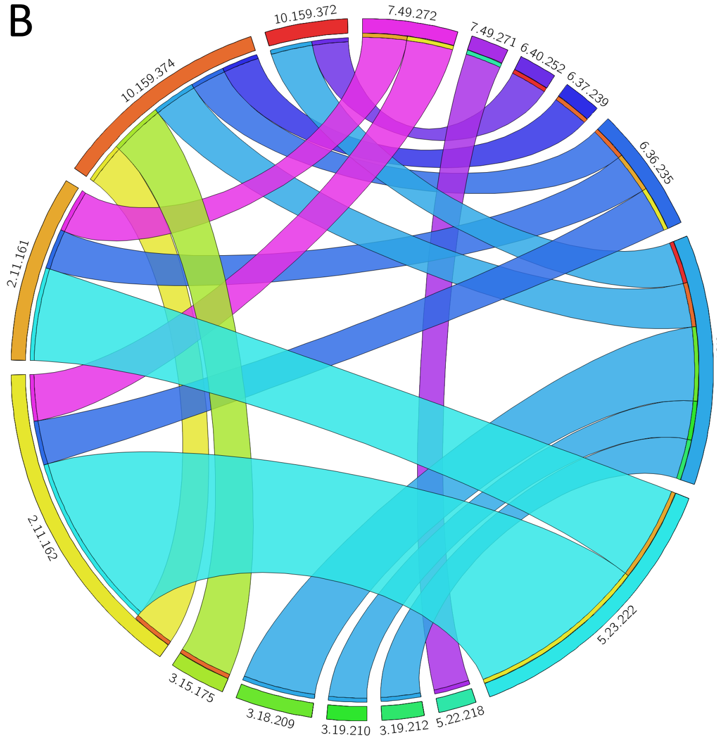


Top unique Type 2 SLE DGCA module pair members

| *lineage* | *enrichment* |
|---|---|
| 10.159.372 | Lug:Platelet. GO:wound.healing |
| 10.159.374 | GO:system.process,protein.heterodimerization.activity |
| **2.11.161** | **Lug:Plasma-Cell. GO:organic.substance.metabolic.process** |
| **2.11.162** | **Lug:Plasma-Cell. GO:response.to.unfolded.protein** |
| 3.15.175 | Lug:Neutrophil. GO:cell.activation.involved.in.immune.response |
| 3.18.209 | GO:cell.diff,polymeric.cytoskeletal.fiber |
| 3.19.210 | GO:GTPase.regulator.activity,interleukin-18-mediated.signaling.pathway |
| 3.19.212 | GO:interleukin-1.receptor.activity,interleukin-1-mediated.signaling.pathway |
| 5.22.218 | Anc:B.cells. GO:cytokinesis |
| **5.23.222** | **Anc:B.cells. GO:extracellular.matrix.organization** |
| 6.36.230 | Lug:Monocyte. GO:response.to.virus |
| 6.36.235 | GO:transferase.activity,transferring.hexosyl.groups,protein.glycosylation |
| 6.37.239 | GO:Rho.GTPase.binding,Rac.GTPase.binding |
| 6.40.252 | GO:inflammatory.response,L-type.voltage-gated.calcium.channel.complex |
| 7.49.271 | GO:stem.cell.population.maintenance,maintenance.of.cell.number |
| 7.49.272 | GO:cytosol,RNA.binding |

**Supplementary figure S7.** Differentially expressed gene pairs distinguish Type 1 & Type 2 SLE patients. Top type 1 SLE and type 2 SLE DGCA intermodular pair totals visualized using circos plots to illustrate the degree of intermodular pair totals between gen3 module largely unique to each condition. Type 1 SLE gene pairs **(A)** and Type 2 SLE gene pairs **(B).** Bolded modules were involved in the highest numbers of totaled module pairings.
