## Supplementary Figure S8 for "The Molecular Endotypes of Type 1 and Type 2 SLE"

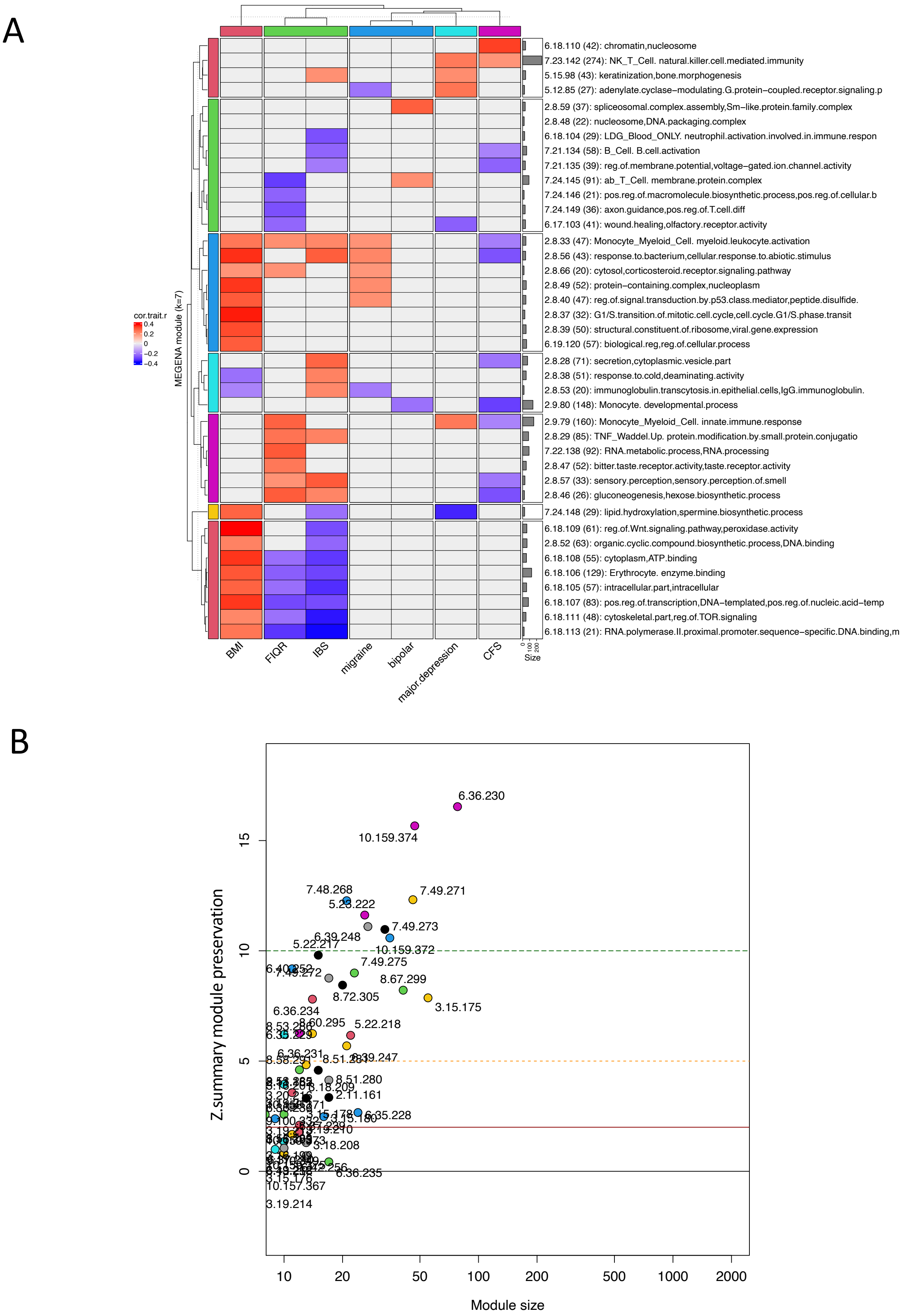


**Supplementary figure S8.** Type 1/2 SLE MEGENA gen3 module preservation amongst the gen3 GSE67311 fibromyalgia (FM) MEGENA modules. Heatmap of the top 40 gen3 MEs significantly (p<0.2) correlated to the seven available patient traits in GSE67311 (**A**). Module preservation composite zsummary (z.summ) statistics calculated using the gen3 type 1/2 SLE modules as reference tested against the gen3 GSE67311 FM modules. Modules are considered minimally preserved with a z.summ >2 (red line), moderately preserved at z.summ >5 (orange line), and well preserved at z.summ >10 (green line) (**B**).
