## Supplementary Figure S9 for "The Molecular Endotypes of Type 1 and Type 2 SLE"

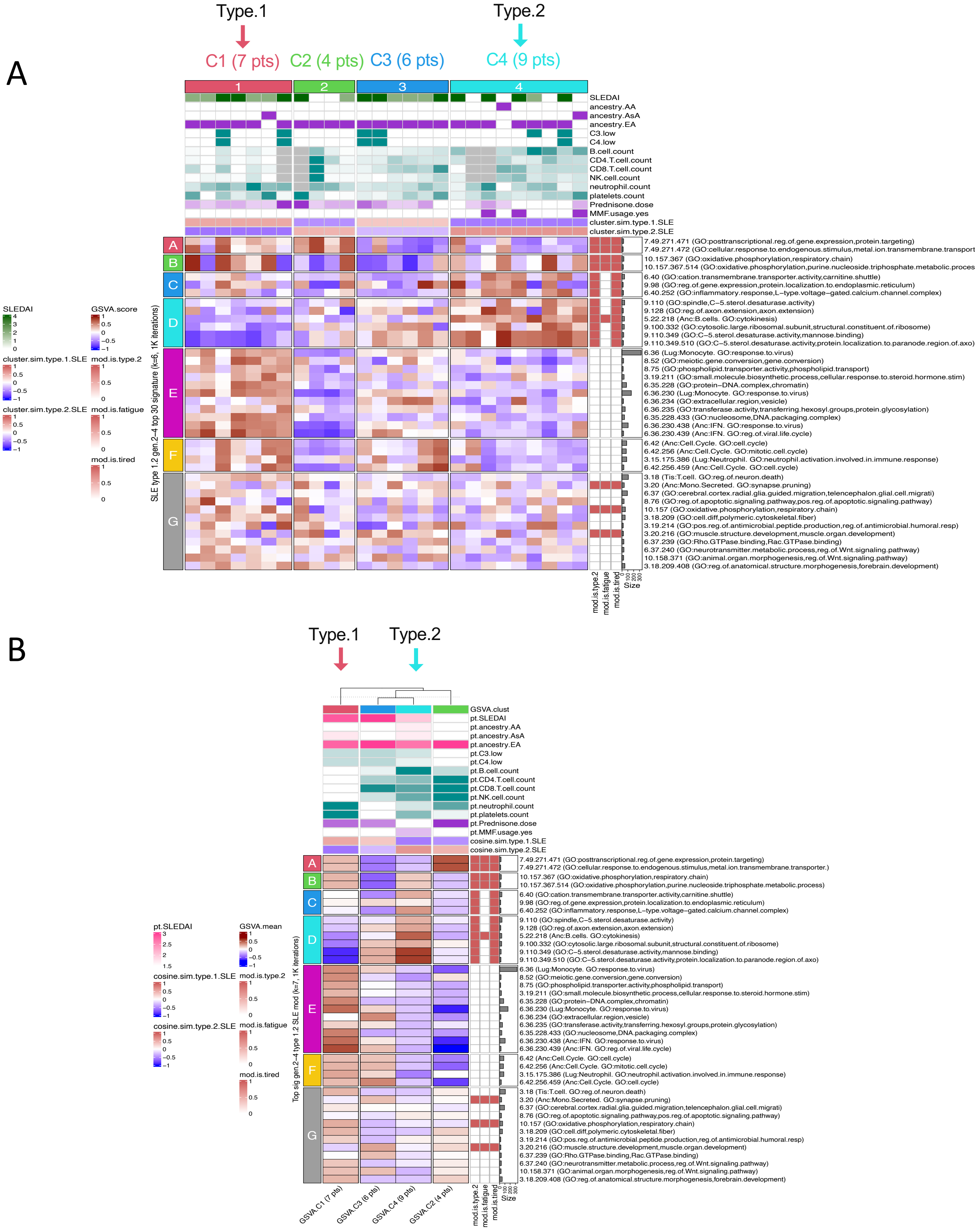


**Supplementary figure S9.** Type 1/2 SLE molecular signatures identify a small subset of 9/26 (34.6%) of subjects with inactive SLE (SLEDAI <6) from GSE49454 exhibiting enrichment of type 2 gene modules. The top5k rowVar genes from GSE49454 were analyzed by GSVA using the top 40 type 1/2 SLE cohort gen.2-4 modules as gene signatures. Column annotations include cohort (healthy or SLE), SLEDAI score and ancestral background (AA African ancestry, AsA Asian ancestry, EA European ancestry, and other), various lab measurements, and mean cluster cosine similarity to bona fide type 1 and type 2 sample results. Inactive SLE patient cluster 1 (7 patients) is most similar to type 1 SLE signatures and inactive SLE patient cluster 4 (9 patients) is most similar to type 2 SLE signatures. Clusters 2 and 3 had only weak similarities to type 1 or type 2 SLE (sim < 0.3). Row annotations indicate modules that were significantly correlated to type 2 SLE, fatigue, and tired. Columns were stably clustered (1k iterations) into k=4 patient clusters and rows optimally clustered into k=7 groups of modules (**A**). GSVA enrichment score row means and sample traits were calculated for the four GSVA patient clusters (**B**).
