## Supplementary Figure S10 for "The Molecular Endotypes of Type 1 and Type 2 SLE"

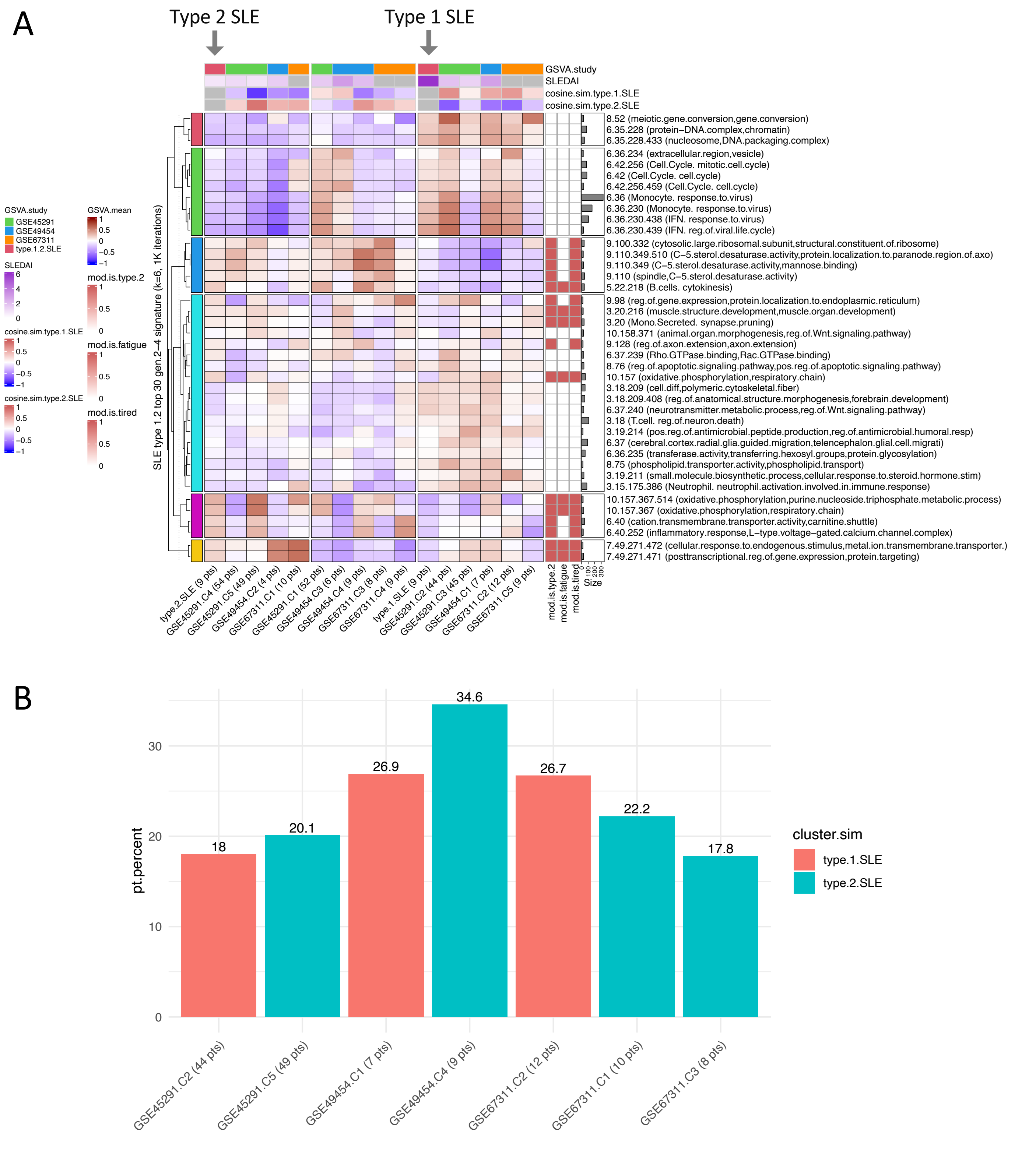


**Supplementary figure S10.** Type 1/2 SLE modules identify subsets of inactive SLE and classic FM patients. Mean GSVA patient cluster enrichment scores were aggregated from GSE45291 and GSE49454 inactive SLE patients, and GSE67311 classic FM patients and visualized together. Patient clusters were further optimally clustered into three meta groups by k means clustering (1k iterations). Column annotations include SLEDAI and cosine similarities to type 1 SLE or type 2 SLE patient clusters (SLEDAI scores were unavailable from the GSE67311 FM study). Row annotations indicate modules that were significantly ($p<0.2$) correlated to type 2 SLE, fatigue, and tired. Arrows indicate the type 1 and type 2 GSVA patient sample profiles that were used as references for cosine similarity testing (**A**). Amongst the patients that significantly (cosine sim > 0.2) resembled type 2 SLE were 49/244 (20.1%) of GSE45291 inactive SLE patients (cluster 5), 9/26 (34.6%) of GSE49454 inactive SLE patients (cluster 4), and 18/45 (40%) of GSE67311 classic FM patients (clusters 1 and 3) (**B**).
