## Supplementary Methods for "The Molecular Endotypes of Type 1 and Type 2 SLE"

**Supplementary Materials & Methods**

**Patient Population**

All patients were adults (≥18 years old) who met 1997 ACR or 2012 SLICC criteria for SLE (1,2). These patients were enrolled in the Duke Lupus Registry (DLR), a prospective registry of adult patients with SLE who receive rheumatology care from six treating rheumatologists in the outpatient clinics of Duke University Medical Center. All patients signed informed consent to participate in the registry; all patients in this analysis provided informed consent to collect RNA samples at clinic visits (Duke Health IRB Pro00008875). This was a cross-sectional analysis on a selected subset of 22 patients (Duke Health IRB Pro00094645), 4 of which were removed from as statistical outliers. On average, patients had been diagnosed with SLE for 15.8 years (SD: 7.3) and 55% had a history of lupus nephritis. Most patients were female (17) with one male. Three ancestral backgrounds were represented including 11 of African ancestry (AA), 6 of European ancestry (EA), and one patient of Hispanic ancestry (HA), and the mean patient age was 41 (**supplementary fig S1.A).** Samples were accompanied by 106 clinical and molecular attributes (supplementary table) notably including SLEDAI, polysymptomatic distress score (PSD), PGA Type 1, PGA Type 1, anti.dsDNA, anti-Smith, anti-La, anti-RNP, and complement C3 assays, and yes/no usage of the immunotherapeutics prednisone, mycophenolate mofetil (MMF), and duloxetine.

**Data Collection**

At routine visits in the DLR Clinic, patients completed the Polysymptomatic Distress Scale (PSD), that includes two subscales: the widespread pain index (WPI) and symptom severity score (SSS) (3–6). For the WPI, patients report the number of areas out of 19 where they have experienced pain in the previous month. For the SSS, patients report the presence and severity of fatigue, cognitive symptoms, and waking unrefreshed over the past month, as well as whether they had experienced a headache, pain or cramps in the lower abdomen, or depression in the last 6 months. The SSS ranges from 0 to 12 and the total is added to the WPI for a total PSD score of 0-31.

In addition to patient-reported measures, patients’ treating rheumatologists completed disease activity measures, including the SLE Disease Activity Index (SLEDAI), Physician’s Global Assessment of Disease Activity (PGA) for Type 1 activity, and a PGA for Type 2 activity (2,7,8) or the PGAs, rheumatologists scored the severity of Type 1 & 2 SLE activity separately on scales from 0 (no activity) to 3 (severe activity). Medications, vital signs, and laboratory measures at each visit were also documented. All data was stored in a secure REDCap database.

**Patient Stratification**

SLE patients were divided into distinct clinical groups based on physician- and patient-reported assessments of Type 1 & Type 2 SLE activity. Type 1 SLE activity was measured by clinical SLEDAI (scored without laboratory measures), full SLEDAI (scored with laboratory measures), presence of active lupus nephritis, and Type 1 PGA. Type 2 SLE activity was measured by Type 2 PGA and patient-reported PSD scores. Patients were divided into classifications of Minimal, Type 1, Type 2, and Mixed based on the extent of their Type 1 & 2 SLE activity (Supplementary Table 6). For this study, 9 patients were identified who had Type 1 and 9 patients with Type 2.

**Gene expression data and gene filtering**

Whole blood was collected in PAXgene Blood RNA tubes. After removal of ribosomal RNA and globin transcripts with the Ribo-Zero Globin Removal kit (Illumina) , stranded libraries were prepared with the TruSeq Library prep kit (Illumina) and hybridized to a flow cell for sequencing with the Illumina HiSeq platform. Raw RNAseq output counts were VST normalized using the R DESeq2 package (9). Genes were further annotated using the R biomaRt (10) library and those without mappings to known proteins were discarded. Duplicate gene symbols were removed using the collapseRows function in the R WGCNA package (11). The top 5,000 row variance (top5k rowVar) genes determined using standard deviation between samples were retained for further analysis.

**Differentially Expressed Gene (DEG) analysis**

The R limma package (12) was used to perform DEG analysis between cohort (type.1.SLE vs type.2.SLE) and calculate empirically Bayesian corrected log fold changes (LFCs). P-values were adjusted for multiple comparisons using Benjamini-Hodgkin adjustment.

**Principal component analysis (PCA) & mixed model patient clustering**

Sample attributes including subjective clinical ascertainments and objective laboratory assay results were encoded as discrete binary values (no=0 or yes=1) or retained as continuous numerical values. The core R prcomp function and PCAtools package (13) was used to conduct exploratory PCA on the top5k rowVar genes. The first four PCs were correlated to the sample attributes using R lm linear regression and the top 20 traits contributing to PC variance visualized using R ComplexHeatmap (14).

**Multiscale Embedded Gene Co-expression Network Analysis (MEGENA)**

The MEGENA (15) R package was used to generate a gene coexpression network by inputting the top5k rowVar genes. MEGENA multi-scale clustering analysis (MCA) formed lineages of gene modules followed by identification of densely intraconnected hub genes using multi-scale hub analysis (MHA). Modules were assigned “lineage” names based on their multiscale pedigree from the root MEGENA module. The prcomp package was utilized to perform singular value decomposition and calculate MEGENA module eigengenes (MEs), equivalent to the first principal component calculated amongst the variance of a given MEGENA module. MEGENA MEs were correlated to the numerically encoded sample traits.

**Coexpressed gene module annotation**

Module gene symbols were overlapped with a number of annotation tools (16), as well as the publicly available Gene Ontology (GO) signatures (17). Annotations of MEGENA modules were considered significant if there were at least 3 overlapping gene symbols between the module gene symbols and annotation signature gene symbols, and the Fisher’s p value statistic of the overlap was p < 0.2. Where there were multiple overlaps, the most significant overlap was assigned. For selection of a given GO annotation, all GO annotations significant by p < 0.2 per the GO enrichment algorithm were ranked in order of decreasing module coverage.

**Coexpression gene network PFN visualization**

The MEGENA planar filtered coexpression network (PFN) of the top5k rowVar genes was imported into Cytoscape along with gene node annotations including functional enrichments, hub node identification, and all generation/scale levels a gene was inherited into. The resulting figure included hub node labels sized according to their scaled degree of intramodular connectedness. The PFN gene nebula was subsequently colored and annotated based on additional relevant information.

**Sample trait intracorrelations**

Correlation of sample traits to the MEs of all relevant MEGENA modules identified 40 significant (p<0.05) correlations. These top 40 sample trait correlations (sig trait corrs) were used as inputs to the R corrplot package (18) to generate a top 40 sample traits intra-correlated correlogram. A bubble plot of the intra-correlations of MEs to patient traits was generated as an alternative representation of the ME correlation plot. The MEs of the top 40 gene expression modules were correlated with various clinical features and the correlation coefficients of the associations plotted on a two-dimensional matrix as described by Zhang, *et al.* (19)

**Coexpression module preservation in GSE67311 Fibromyalgia**

Gene expression data from fibromyalgia patients was obtained from the Gene Expression Omnibus (GEO) study GSE67311 (20). This study originally included 70 fibromyalgia patients and 70 matched controls. The raw files from the Affymetrix® Human Gene 1.1 ST Peg arrays were RMA normalized using the R affycoretools package (21). COMBAT batch correction was applied using the R SVA package (22) followed by normalization to commonly known house-keeping (HK) genes. The normalized top5k rowVar genes from fibromyalgia patients were submitted to MEGENA for formation and annotation of gene coexpression modules. We calculated module preservations between the SLE type 1/2 and GSE67311 fibromyalgia patients MEGENA modules utilizing an algorithm that generates z.summ composite scores of 20 preservation metrics (11).

**Coexpression module correlation and enrichment plots**

Sunburst correlation plots were generated using the R plotly (23) package to illustrate MEGENA significant (p < 0.05) ME correlations to demographics and clinical features. These were followed by significant ME correlations to patient type (type.1.SLE or type.2.SLE), full (anti.dsDNA validated) SLEDAI and PSD score. Enrichment sunbursts were generated by statistically overlapping the gene symbols within a given MEGENA module with the various enrichment lists previously mentioned. An overlap was significant if there were at least 4 gene symbols overlapping with an enrichment signature and the Fisher’s p.val of that overlap was < 0.2.

A heatmap was generated using ComplexHeatmap visualizing the top 40 sample trait correlations to the 40 MEGENA modules that were significantly (p<0.2) correlated to cohort (type.1.SLE=0 and type.2.SLE=1). Module gene symbols were used to programmatically query the STRING database (24) and calculate the percentage of genes within a given module predicted to have known protein-protein interactions (PPI) ranging from 0 to 100%.

**MEGENA module eigenegene (ME) correlations to patient gene expression**

The MEs of the 40 significant modules were correlated to mean gene expression of a given module per patient and visualized using Complex heatmap. Columns of patients were clustered using idealized k-means clustering. Rows were annotated in a manner similar to the trait correlations heatmap and included STRING PPI intraconnectedness and module preservation with GSE67311 fibromyalgia patient samples. Mean ME correlations and patient traits per patient cluster were calculated and visualized as a complex heatmap.

**Gene Set Variation Analysis (GSVA)**

The GSVA (25) (V1.25.0) R software package was used as a non-parametric, unsupervised method for estimating the variation of pre-defined gene sets over all MEGENA module log2 gene expression values. Input genes were employed only if the interquartile range (IQR) of their expression across the samples was greater than 0. Enrichment scores (GSVA scores) were calculated non-parametrically using a Kolmogorov Smirnoff (KS)-like random walk statistic. The enrichment scores (ES) were the largest positive and negative random walk deviations from zero, respectively, for a particular sample amongst the module gene set. The GSVA scores were used an input for unsupervised stable k-means clustering, and two different disease phenotypes or clusters were identified. Mean GSVA enrichment scores and patient traits per patient cluster were calculated and visualized as a complex heatmap.

**Differential Gene Co-Expression Analysis (DGCA)**

The R DGCA (26) software package was utilized to identify differentially expressed gene pairs between type.1.SLE & type.2.SLE patients. Significant DGCA pairs were queried against the CellTalk (27) repository of 3,398 human ligands and receptors. The plotly package was utilized to generate sunbursts of the totaled DGCA intermodular pairs between the top unique interconnected gen3 modules and the modules labeled with their top functional annotation. Cytoscape was used to visualize the intramodular and intermodular connections/edges found between various interconnected gen3 MEGENA modules.

**Patient Age Adjustment Analysis**

Type 1 SLE patients 168 and 251, and type 2 SLE patients 028 and 230 were removed from the data set leaving 7 patients from each cohort with balanced age distributions. Gene expression of the 14 patients were submitted to MEGENA analysis forming new modules and the top 40 type 2 SLE ME module correlations visualized as a complex heat map. GSVA was performed on the top5k rowVar gene expressions using the top 40 age-balanced modules as signatures.

The top5k rowVar gene expressions of the original 18 patients were adjusted using linear modeling with age as a covariant. These were submitted to MEGENA analysis forming new modules and the top 40 type 2 SLE ME module correlations visualized as a complex heat map. GSVA was performed on the top5k rowVar gene expressions using the top 40 age-adjusted modules as signatures.

**Inactive SLE Data Sets Analysis**

The top 5,000 row variance genes from inactive lupus studies (SLEDAI<6) GSE45291 and GSE49454 were used submitted to GSVA analysis and the GSVA enrichment scores visualized in the manner previously described. Mean GSVA enrichment scores and patient traits per patient cluster were calculated. The mean scores per patient cluster underwent cosine similarity tests using the R lsa package (28) against the two type 1/2 SLE mean GSVA patient clusters. visualized as complex heatmaps. Column annotations included patient traits from their respective studies along with cosine similarity scores ranging from -1 to +1.

**Active SLE Data Set Analysis**

The top 5,000 row variance genes from active lupus study GSE88884 (Illuminate-2) were used submitted to GSVA analysis and the GSVA enrichment scores visualized in the manner previously described. Mean GSVA enrichment scores and patient traits per patient cluster were calculated. The mean scores per patient cluster underwent cosine similarity tests to the Type 1/2 SLE GSVA means reference clusters and visualized in a manner similar to the inactive SLE GSVA means heatmaps. The differences between the proportions of mild and severe pain and fatigue groups in each k-means cluster were tested using the R stat package proportion test. The distribution of mild and severe fatigue and pain groups in each cluster were visualized using bar plots. Patient clusters marked as (*) exhibited a significant difference between the frequency of severe and mild fatigue or pain, respectively.

**Aggregation of Type 1/2 SLE, Inactive SLE, Active SLE, and Classic FM GSVA Means Clusters**

The GSVA mean enrichment scores from the four studies were aggregated into a single matrix, clustered using idealized k-means, and visualized using a complex heatmap. Column annotations included SLEDAI (where available), and cosine similarity to the Type 1/2 SLE reference clusters. Row annotations included module correlation to Type 2 SLE, “fatigue”, and “tired”. Bar plots were generated indicating the percent of patients in the inactive SLE, active SLE, and classic FM patients that significantly resembled Type 1 or Type 2 SLE per cosine similarity.
